# CoWWAn: Model-based assessment of COVID-19 epidemic dynamics by wastewater analysis

**DOI:** 10.1101/2021.10.15.21265059

**Authors:** Daniele Proverbio, Françoise Kemp, Stefano Magni, Leslie Ogorzaly, Henry-Michel Cauchie, Jorge Gonçalves, Alexander Skupin, Atte Aalto

## Abstract

We present COVID-19 Wastewater Analyser (CoWWAn) to reconstruct the epidemic dynamics from SARS-CoV-2 viral load in wastewater. As demonstrated for various regions and sampling protocols, this mechanistic model-based approach quantifies the case numbers, provides epidemic indicators and accurately infers future epidemic trends. In situations of reduced testing capacity, analysing wastewater data with CoWWAn is a robust and cost-effective alternative for real-time surveillance of local COVID-19 dynamics.

---

Effective mitigation of the COVID-19 epidemics relies on reliable estimates of the epidemic dynamics. Analysing SARS-CoV-2 abundance in wastewater offers a cost-effective alternative to population-based large scale testing [1, 2] and is largely independent of healthcare-seeking behaviors, access to clinical testing and asymptomatic cases [3]. It thus bears the potential for faster and more reliable early warning indications for long-term epidemic surveillance [4, 5, 6]. To date, more than 50 countries and 260 universities have wastewater surveillance systems in place [7]. However, despite improved experimental procedures and data processing [8], most of the current analysis approaches are restricted to qualitative and semi-quantitative retrospective studies of lagged correlations [9, 10]. These have limitations in quantitatively inferring the shedding population or in providing reliable projections of the epidemic dynamics. To address this challenge and fully exploit the potential of SARS-CoV-2 wastewater abundance measurements, we developed CoWWAn as an automated approach that causally infers the shedding population, estimates the effective reproduction number *R*_eff_, and provides projections of future epidemic trends. Quantifying these variables allows assessing the epidemic status within a region and comparing it between regions, and supports effective mitigation policy making.

CoWWAn couples a mechanistic epidemiological model, describing the infection dynamics through a Susceptible-Exposed-Infectious-Removed (SEIR) process [11], with an Extended Kalman filter (EKF) [12] (Fig. 1a) for robust integration of noisy measurement data into a predictive modelling framework (Methods). The underlying SEIR model allows interpreting the inferred infection dynamics in terms of transmitting interactions, overcomes the interpretability and extrapolation limitations of correlation-based statistical approaches [13, 14] and, once calibrated, provides reliable estimates of the shedding population and future development of the epidemic. To demonstrate its general applicability, we applied CoWWAn to public datasets from 12 regional areas from Europe and North America (Supplementary Tab. 1), associated with different population sizes and based on different wastewater data processing protocols. Details on datasets, list of considered regions and selection criteria are given in Methods and Supplementary Tab. 1, Supplementary Figs. 1 and 2.

**Figure. 1.**
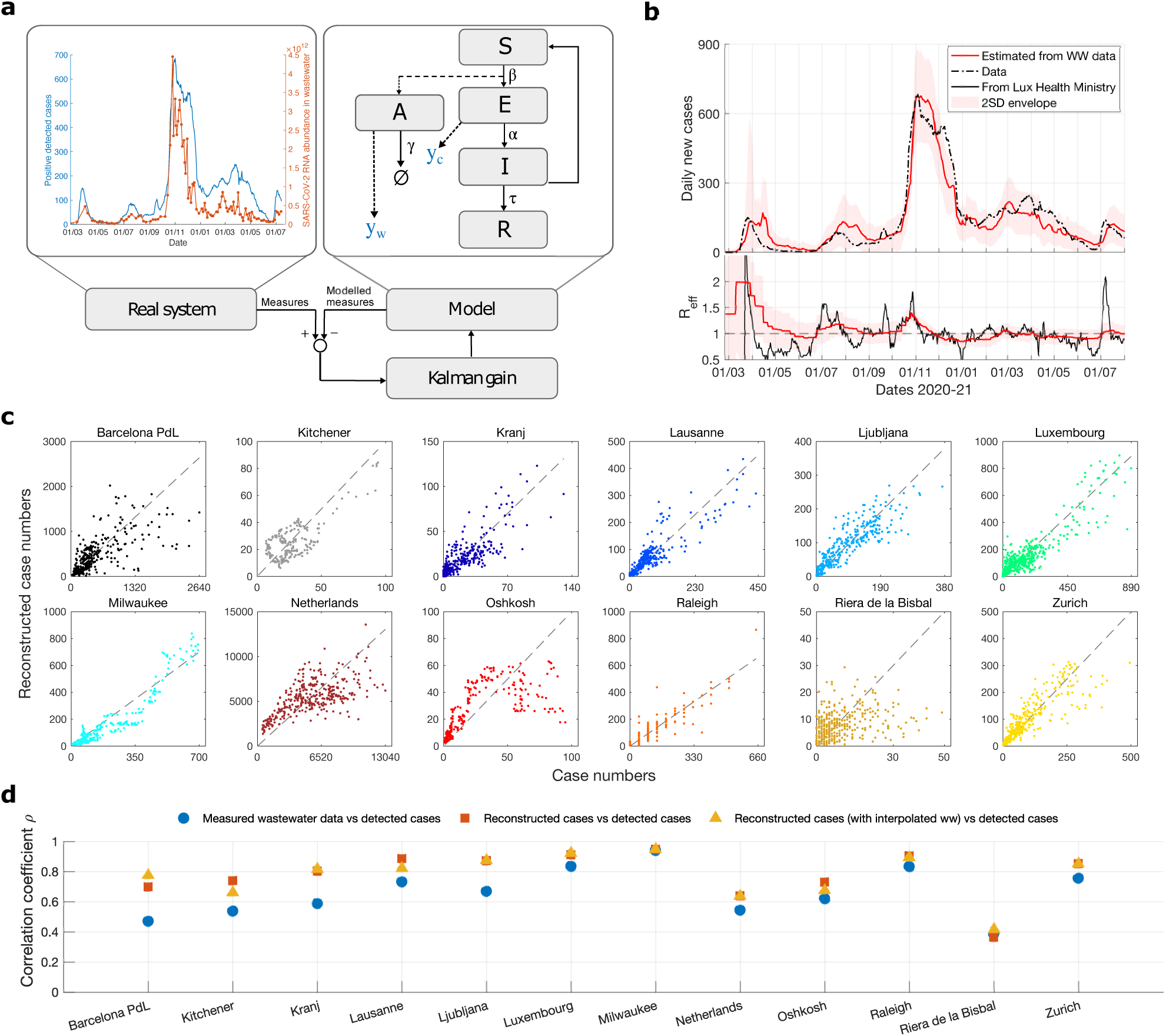
CoWWAn: a mechanistic model-based approach for reliable inference of the COVID-19 dynamics from SARS-CoV-2 viral load in wastewater. **a**, The CoWWAn approach combines an epidemiological SEIR model – complemented with a compartment for active cases producing virions to wastewater – with an Extended Kalman filter for robust estimates of daily new cases from wastewater abundance data. **b**, Reconstruction example for Luxembourg. Top: Comparison of case numbers, detected (black line) or reconstructed by CoWWAn from wastewater data (red), including the 2 Standard Deviations ≃ 95% confidence interval (shadowed region). Bottom: *R*_eff_, estimated by CoWWAn (red, with its associated 2 SD shadowed region) or officially reported by the Luxembourg Ministry of Health. **c**, Reconstruction results for all considered regional areas, compared with detected case numbers. The dashed line represents equal values. **d**, Pearson’s correlation coefficients *ρ* from linear regression between detected cases and measured wastewater data (blue), *ρ* between detected cases and CoWWAn-reconstructed case numbers from wastewater data (red, corresponding to correlation values from panels c), and *ρ* between CoWWAn-reconstructed case numbers from wastewater data (after interpolating wastewater data) and detected cases (yellow).

After appropriate calibration to test cases, CoWWAn quantitatively reconstructs the time evolution of observed cases from wastewater data (Fig. 1b) by inferring the internal variables and parameters of the SEIR model. These include the susceptible, exposed and infectious population fractions, daily detected cases and time-dependent infection rate (Methods). In our case studies, full time series data were used for calibration for each region. When clear regime shifts in testing/sampling protocols are observed, the model can be re-calibrated appropriately to improve the performance, like for Kitchener (Methods and Supplementary Tab. 2). To infer the global shedding population, the model needs additional information on the ratio of total and detected cases, typically obtained from prevalence studies (Methods). A comparison with linear regression (after data curation to reduce the noise) reveals that CoWWAn’s inferences achieve consistently higher correlation (Fig. 1d, blue and red sets), demonstrating the power of our mechanistic-based approach. These observations hold for all considered regions (Fig. 1c and Supplementary Fig. 3-14): the correlation coefficient *ρ* between inferred case numbers and true detected case numbers is typically in the range between 0.7 and 0.9 even for rather noisy data like Netherlands. Frequent sampling improves the model calibration and the subsequent reconstruction performance, like for Luxembourg with *ρ* = 0.91 for two probes/week and Milwaukee with *ρ* = 0.95 for two (sometimes more) probes/week compared e.g. to Barcelona with *ρ* = 0.70 with one probe/week (Fig. 1d). The main discrepancies originate from either unnoticed changes in the share of detected cases or from changes in testing/sampling strategies (Supplementary Fig. 3-14). Detecting such discrepancies can provide additional evidence about potential undertesting and could guide targeted scaling of population tests. Interpolating wastewater data points before the EKF estimation can improve the reconstruction (Fig. 1d, red and yellow sets), in particular for regions with low sampling frequency like for Barcelona Prat de Llobregat (PdL) and Kranj. In general, the Extended Kalman filter improves its predictions as new data points are available, so an adequate sampling rate is recommended to improve its performance.

In addition, CoWWAn estimates the effective reproduction number *R*_eff_, an essential indicator for the trends of epidemic diffusion in a community [15], which depends on containment measures, infectivity of viral variants, population behavior and other factors. As exemplified for Luxembourg (Fig. 1b), the *R*_eff_ values inferred by CoWWAn from wastewater data are consistent with the indicator reported by the Ministry of Health on its website (Methods) and exhibit the same noteworthy trends: the three waves in 2020 (March, June and late October), a small rebound in March 2021 and one wave in late June 2021, all characterised by *R*_eff_ > 1. For all other considered regions as well, wastewater-based *R*_eff_ values are consistent with those estimated from case numbers (Supplementary Fig. 3-14) and are usually smoother due to sampling frequency and independence to testing schemes.

CoWWAn’s underlying SEIR model permits mechanistic-based projections of the infection dynamics, for effective monitoring of the epidemic. To produce projections of future trends, it is possible to stop the reconstruction at any desired time and simulate the model forward, starting from the latest state estimate and keeping the transmission parameter constant (Methods). For the epidemic dynamics in Luxembourg, Fig. 2a shows an example of such 7-days projections for each day of wastewater sampling, where the number of detected cases (blue) is compared with the projected numbers derived from wastewater data or from case number data. Wastewater-based projections are well correlated both with case-based projections (*ρ* = 0.95) and with true case numbers (*ρ* = 0.94). Overall, for the different epidemic phases and all considered regions, the projections compare well with the real case data and with the case-based projections (Supplementary Fig. 3-14). To quantify the projection performance, we determined the average standardised projection error as the average discrepancy between projected and actual case numbers in the corresponding time frame, normalised to case numbers and equivalent population (Methods Eq. 10). The performance of our wastewater-based pipelines is usually slightly lower, as they reconstruct the case numbers themselves before making the projections, but remains similar with that of case-based projections: all regional estimates lie within one standard deviation of the 1:1 (equal performance) line (Fig. 2b). The only exceptions are projections for Oshkosh, probably due to under-testing during late 2020 (Supplementary Fig. 2) which induced discrepancies in the detected cases fraction, and Kranj, whose low case numbers are subject to larger uncertainties (Supplementary Fig. 5). In general, the largest discrepancies are observed when case numbers plateau or decline after a rapid increase, yielding a potential overshoot of the projections (Fig. 1a and Supplementary Fig 3-14). This effect is associated to large changes in social activities during epidemic waves and rapid implementations of stricter restrictions, which are not explicitly included in the model but implicitly learned from the epidemic curve by the EKF with some delay.

**Figure. 2.**
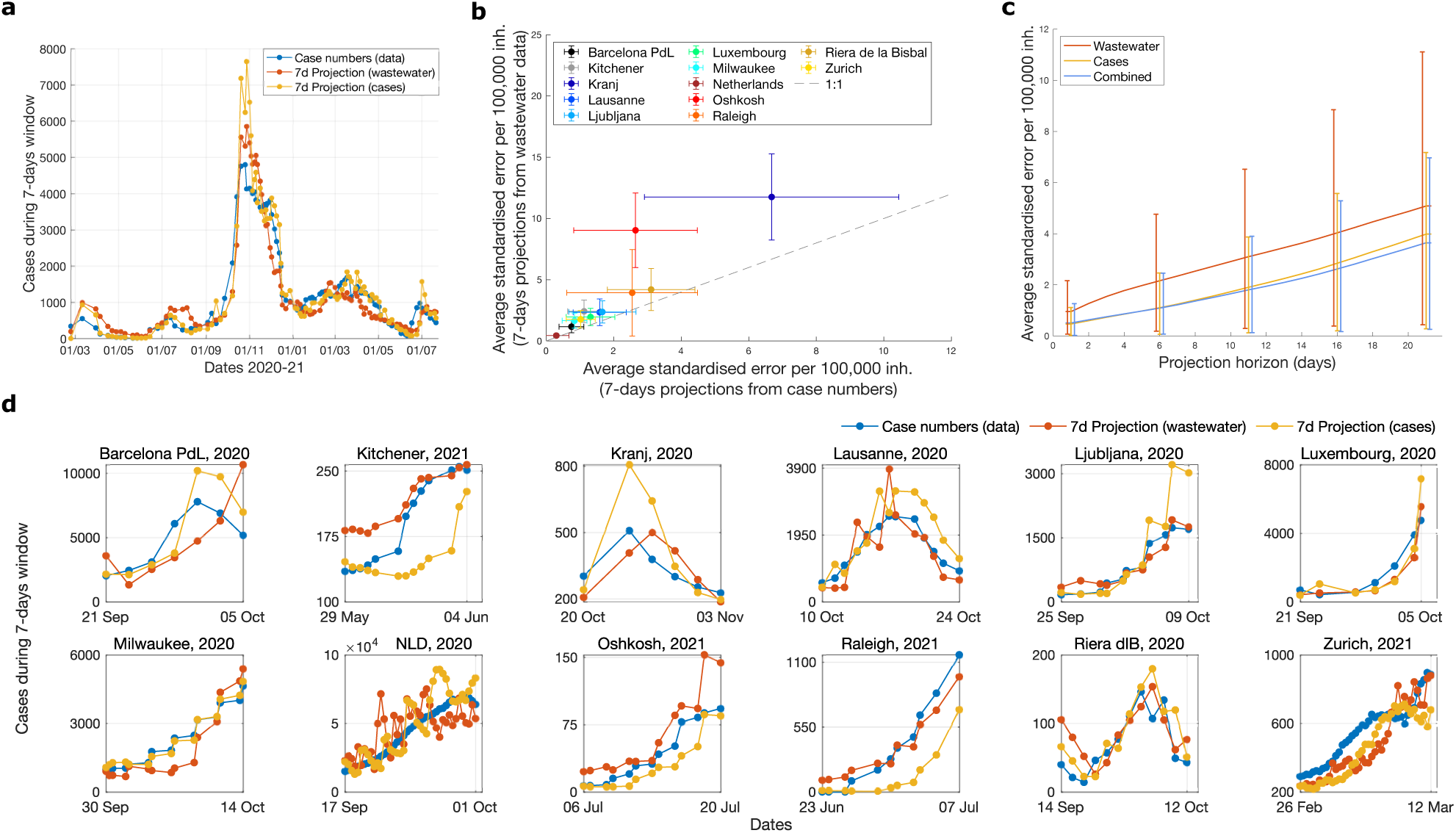
Projections of future epidemic trends using CoWwAn. **a**, Projection examples for Luxembourg, comparing projections over the 7-days ahead of each point (either estimated from case numbers or wastewater data) with the true detected cases in the same time period. **b**, Comparison of wastewater-based projections with cases-based projections. The performance is evaluated in terms of average standardised error, normalised to equivalent population. The dashed line represents equal values. Error bars correspond to one standard deviation. **c**, Projection performance for different projection horizons (mean and 80th percentiles over the considered regions; outputs for single countries in Supplementary Fig. 17) for three CoWWAn’s inputs: case numbers, wastewater data, or both data combined. **d**, Short-term projections used to identify robust trends in epidemic resurgence, for different examples (one per region; other examples in Supplementary Fig. 16). We compare 7-days projections from case numbers and from wastewater data with the true detected case numbers. For all panels, “inh.” stands for inhabitants.

The standardised error grows quite linearly with increasingly long projection horizons (Fig. 2c), where wastewater projections are more stable (their uncertainty grows slower for longer projection horizons) than those based on case numbers as they are usually less susceptible to daily fluctuations (Supplementary Tab. 2). Due to heterogeneous and evolving adaptations of population behavior and institutional measures, epidemic forecasts are typically only meaningful for relatively short time horizons [16]. In particular for the real-time detection of impending epidemic resurgence, distinguishing between fluctuations and robust increases is crucial to optimise the true positive signals and minimise the false negatives. CoWWAn addresses this challenge by the EKF-based projections, which capture robust trends in the epidemic dynamics and allow for early warning of COVID-19 resurgence from case-based and wastewater-based projections (Fig. 2d and Supplementary Fig. 16). On the other hand, long-term projections that assume no changes in infection dynamics can be useful for counterfactual analysis of current measures (Supplementary Fig. 15). This analysis demonstrates the potential of wastewater data to inform investigations of incoming trends and quantifies the precision of this cost-effective surveillance method. Finally, CoWWAn’s EKF-based approach enables integrating different types of data to further improve the quality of projections. Including both wastewater and case data slightly but systematically improves the projection accuracy compared to case data alone (Fig. 2c and Supplementary Tab. 2), suggesting that wastewater data contains independent information about the state of the epidemics [17].

In summary, leveraging wastewater data with CoWWAn as an automated and mechanistic approach allows for new avenues for epidemic monitoring. In situations of reduced population testing, CoWWAn can support the reconstruction of the infection curves from wastewater data and allows projections of future trends, in particular close to epidemic resurgence. Hence, it can trigger community-wide alerts to elicit targeted studies. Since hospital admission is typically downstream of the susceptible-exposed-infectious flow [18], an early detection of positive increases, supported by quantitative models that account for noise, could provide crucial information for healthcare management [19, 20]. The flexibility of our freely available approach, its ease of implementation and its performance make it an important tool for long-term monitoring and support of epidemic mitigation.

## Data Availability

Data produced in the study is available together with the code. Other used datasets are available online. Website addresses for different datasets can be found in Supplementary table 1.

https://gitlab.lcsb.uni.lu/SCG/cowwan

## Author contributions

D.P. and A.A. conceptualised the project. L.O. and H.M.C generated the data for Luxembourg. D.P., F.K., L.O., A.S. and A.A. analysed the data. D.P., A.A. and F.K. designed and developed the model. D.P. and A.A. implemented the code. All authors analysed and interpreted the results. J.G. and A.S. supervised the project. L.O., H.M.C. J.G. and A.S. acquired the funding. D.P. and A.A. wrote the first draft. All authors contributed to and approved the final manuscript.

## Acknowledgments

The authors want to thank the Research Luxembourg COVID-19 Task Force for general support and collaborative spirit. D.P. and S.M. are supported by the Luxembourg National Research Fund (FNR) through PRIDE15/10907093/CriTiCS and F.K. by the FNR project PRIDE17/12244779/PARK-QC. A.A. is supported by the FNR through CORE19/13684479/DynCell. L.O. and H.M.C. are supported by the FNR through the COVID-19-FT2/14806023 /Coronastep+. J.G. is partly supported by the 111 Project on Computational Intelligence and Intelligent Control, ref B18024.

## Competing interests

The authors declare no competing interests.

## Data availability

The wastewater and case numbers data that support the findings of this study are available from the websites listed in Methods (Data) and Supplementary Tab. 1. Luxembourg data for this study are available at gitlab.lcsb.uni.lu/SCG/cowwan.

## Code availability

CoWWAn’s implementation for Matlab 2019b is available at gitlab.lcsb.uni.lu/SCG/cowwan.

## Methods

## 1 Methods

### 1.1 Data

Wastewater data and case numbers were obtained from different sources, listed in Supplementary Tab. 1 along with the countries considered and their associated equivalent population (*i*.*e*., the ratio of the sum of the pollution load collected during 24 hours by sewage facilities and services to the individual pollution load in household sewage produced by one person in the same time. It is a proxy of the number of people who contributed to the wastewater load). Our pipeline was tested on datasets with different normalisation protocols for wastewater data, to show its general applicability after proper calibration. We refer to each source for details about the experimental protocols. The units of measure of wastewater data, according to the official sources, are reported in Supplementary Tab. 1.

The selection criteria for data collection are the following. First, we employed the COVID19 Poops Dashboard [21] to list all worldwide resources about wastewater sampling projects; among those, we focused on those having readily accessible databases. To allow proper calibration of the model, we selected time series data starting no later than beginning 2021, having at least one sample per week on average and having the corresponding case numbers available. We rejected wastewater data with smoothing among data points to avoid introducing bias and breaking the causality of projections. Finally, if time series from multiple treatment plants were available from a single regional database, we selected the two representative ones, usually with the largest population.

The selected datasets, from specific wastewater treatment plants or covering bigger regional areas, are the following: Barcelona Prat de Llobregat (Spain), Kitchener (Canada), Kranj (Slovenia), Lausanne (Switzerland), Ljubljana (Slovenia), Luxembourg, Milwaukee (USA), Netherlands, Oshkosh (US), Raleigh (US), Riera de la Bisbal (Spain), Zurich (Switzerland). Data from Luxembourg sewage sampling were made available by the Research Luxembourg COVID-19 initiative CORONASTEP (researchluxembourg.lu/coronastep), while case numbers and *R*_eff_ were obtained from the Luxembourg Ministry of Health website (COVID19.public.lu/fr/graph). Other datasets were downoaded from publicly available official sources, listed in Supplementary Tab. 1. All datasets are updated up to August 2021.

Among the data collected, there are some peculiarities. First, Raleigh county reported case numbers normalised to 10,000 inhabitants and rounded to an integer value; their subsequent up-scaling induces a further uncertainty. Second, the countrywide wastewater data for Netherlands are reported as averages over a week. To improve the temporal resolution of the data, we used instead the data from all communal treatment plants, averaging over samples from the same day. Third, the wastewater data from Kitchener have a sudden jump on May 17, 2021 during a time when case numbers remain stable (Supplementary Fig. 1). Interestingly, the performance of our method increased considerably after scaling data after that date by a factor of 0.4, suggesting possible sudden changes in testing strategies. This extra analysis shows the impact of including corrections for different testing policies. Results in the main text are shown without this scaling, but we report results with and without scaling in Supplementary Tab. 2.

A preliminary analysis was carried out to investigate the most prominent features of case numbers and wastewater data, and to inform the development of the model. Considered time series of tested positive case numbers and of RT-qPCR wastewater data, as well as their mutual relationship, are shown in Supplementary Figs. 1 and 2. The figures highlight the close but not perfect correlation between case numbers and wastewater data, stressing both the usefulness of wastewater data for epidemic monitoring, and the importance of models based on complex epidemiological dynamics. The fact that the mutual relationship between case numbers and wastewater data is not perfectly linear justifies the inclusion of a scaling parameter in the cost function used for parameter fitting (Eq. 10 and corresponding paragraph).

### 1.2 The SEIR stochastic model

As a basis for the Extended Kalman filter to model the epidemic dynamics, we use a SEIR model, which has been shown to accurately describe COVID-19 epidemic dynamics [22, 23]. As we aim at estimating community prevalence from noisy data, we choose a simple and descriptive model rather than a complex one, which is difficult to calibrate and could suffer from identifiability issues [24, 25].

The classic, deterministic SEIR model considers Susceptible *S*(*t*), Exposed *E*(*t*), Infectious *I*(*t*) and Removed *R*(*t*) compartments, and population flows governed by rate parameters. The total community population is conserved, i.e. *S*(*t*)+ *E*(*t*)+ *I*(*t*)+ *R*(*t*) = *N* (with constant *N*). To model measurement uncertainties, as well as intrinsic stochasticity in transmission processes and viral shedding, we employ a stochastic version of this SEIR model, associating each transition between compartments with a random process. The SEIR model is based on the assumption that each susceptible person has probability *β*(*t*)*I*(*t*)*/N dt* to become infected on an infinitesimal time interval [*t, t* + *dt*), and that infection events are independent. The number of new infections at [*t, t* + *dt*) is then a random variable from the binomial distribution ℬ (*n, p*) with *n* = *S*(*t*) and *p* = *β*(*t*)*I*(*t*)*/N dt*. Assuming high enough number of cases and stationary rate parameters over a time interval Δ*t* = 1 day [26], the binomial distribution can be well approximated by the normal distribution with mean *β(t*)*S*(*t*)*I*(*t*)*/N dt*, and variance *β*(*t*)*I*(*t*)*/N dt* (1 − *β* (*t*)*I*(*t*)*/N dt*)*S*(*t*)= *β*(*t*)*S*(*t*)*I*(*t*)*/N dt* + 𝒪(*dt*^2^). The same steps can be repeated for all other transitions between compartments. The stochastic SEIR model is then:

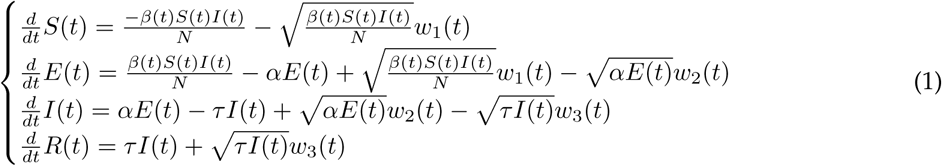

where *w*_*j*_ are mutually independent white noise processes. The *β*-parameter is assumed to be timevarying, reflecting changes in social interaction, other mitigation measures (masks, vaccines, etc.), and varying infectivity of emerging viral variants. *β* will as well be estimated by the Kalman filter.

In order to model viral flows into wastewater, we introduce another variable *A*(*t*) to model the number of active shedding cases producing virions to wastewater. Similarly to above, we incorporate a stochastic processes. The dynamics of *A* is given by:

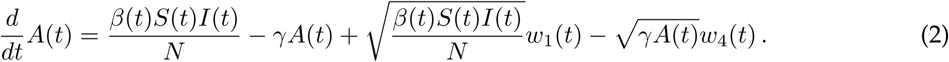

Note that *A* compartment is parallel to *E, I*, and *R*, that is, it still holds that *S*(*t*)+ *E*(*t*)+ *I*(*t*)+ *R*(*t*)= *N*. The influx to the *A* compartment is the same as that to the *E* compartment, while the outflux lumps together the dynamics of viral production [27] and the decay rate of SARS-CoV-2 RNA in water [28, 29]. We do not take into account delays associated with in-sewer travel time, as it was estimated to be significantly lower than the transmission time scales (median of 3.3h [30] versus 1 day). Together, Eq. 1 and Eq. 2 form the combined SEIR-WW system.

The outputs from the model that are compared to the real-world measurements are the number of daily detected cases and the virion abundance in wastewater. The number of detected cases on day *t* ∈ ℕ is assumed to be a share of people passing the incubation period on that day, that is,

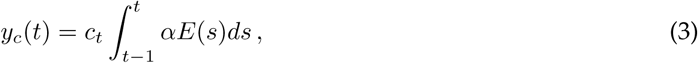

where *c*_*t*_ ∈ [0, 1] is the share of detected cases out of all cases, to account for under-testing and asymptomatic cases (see Model Parameters and Eq. 8 for further discussion). *c*_*t*_ might depend on the day of the week, since there often are some weekday-dependent fluctuations in testing. The virion abundance in wastewater, is assumed to be linearly dependent on *A*,

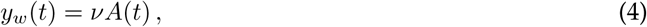

where *ν* is a tuning parameter to reflect the incubation, production and shedding of viral load from infected people [27, 31, 32]. We do not consider explicit corrections linked to precipitations or other environmental factors, as previous studies evaluated them to be poorly correlated with RT-qPCR observations [33, 34]. An implicit tuning is nonetheless included in the fitting, *cf*. Eq. 10.

### 1.3 The complete SEIR-WW-EKF model

In a broad sense, our proposed Kalman filter combines a model of a dynamical system with measurements obtained from the real system that is being modelled. At each time step, the EKF first predicts the next state - the set of all variables - by propagating the old state estimate using the underlying model. From the predicted state estimate, the predicted measurement is calculated using the measurement model. Finally, the state estimate is updated based on the discrepancy of the true measurement and the model-predicted measurement. The model’s state estimate then reflects the state of the real system, and it can be used to predict the system’s dynamics in the future.

In discrete time, which is appropriate to represent the sampling rates, an Extended Kalman filter requires an underlying dynamical model (such as a SIR-like one), its output and associated covariance matrix, and measurement data.

To embed the SEIR dynamical system in the Extended Kalman filter, we formulate a time-discretised state-space version of the dynamical system Eq. 1 by explicit Euler method:

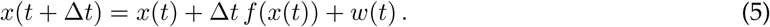

To obtain the number of daily new infections from the model on a given day, an additional auxiliary state variable *D*(*t*) is defined, whose dynamics is given by

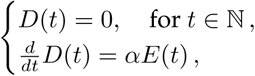

that is, *D(t)* is the difference counterpart of *y*_*c*_*(t)* and is reset every day to keep track of new infections on the current day.

Including the auxiliary variable, the state space is 6-dimensional with variables *x*_*1*…6_*(t)*= [*S, E, I, A, D, β*]*(t)*. Due to conservation of *N, R(t)* is redundant and is therefore omitted. Eq. 5 is complemented with the resetting of *x*_*5*_*(t)* to zero once per day. The function *f(x)* can be represented by a reaction function *r(x)* which is multiplied by the stoichiometric matrix *B*:

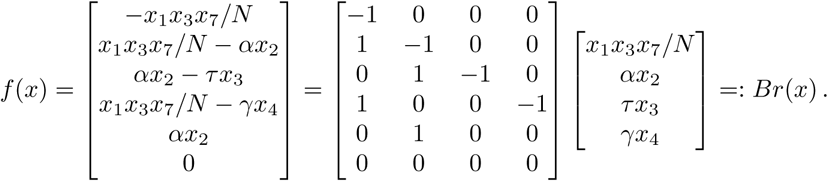

As argued in the previous section, the state noise *w*(*t*) can be well approximated as normally distributed with mean zero and covariance

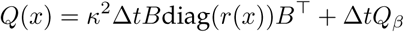

arising from the stochastic model Eq. 1; note that each white noise process *w*_*j*_ for *j* = 1, …, 4 in (1) corresponds to its respective reaction *r*_*j*_*(x)*. The coefficient *k* is used to account for modelling errors. In particular, the SEIR model implicitly assumes a homogeneous and perfectly mixed population. This assumption leads to a rather small uncertainty. The coefficient *k* can also be interpreted as a sensitivity tuning parameter. Lower *k* leads to higher sensitivity but noisy estimates. Higher *k* decreases sensitivity but increases robustness against noise. The parameter *β* has no dynamics through *f*(*x*), but it is updated by the Kalman filter. The matrix *Q*_*β*_ is otherwise zero, except for the element (6,6) being *q*_*β*_, which acts as a tuning parameter controlling the magnitude of change of *β(t)* in one day.

The measurements from the model are either detected cases on a given day and/or wastewater sampling. To this end, we define possible observation matrices:

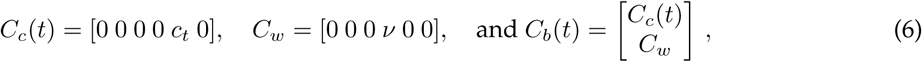

where the sub-indices refer to cases *(c)*, wastewater *(w)*, and both *(b)*. We recall that *c*_*t*_ is the share of detected cases on a day *t*. It is a coefficient that reflects the testing strategy, which often depends on the day (reduced testing on weekends and on public holidays).

The empirical measurements are assumed to be noisy, with an additive, normally distributed noise with mean zero and covariance *U(t)* = diag(*U*_*c*_*(t), U*_*w*_) (or just *U(t)* = *U*_*c*_*(t)* or *U(t)* = *U*_*w*_ if only one of the measurements is available). The variance of observed cases, *U*_*c*_*(t)*, is obtained by assuming that cases are detected independently with probability *c*_*t*_. This leads again to a Binomial distribution for detected cases with mean *c*_*t*_*D(t)*, where *D(t)* is the number of new infections on day *t*. This is unknown to us, and we use a smoothed estimate 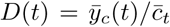 (barred variables stand for 7-days moving averages). The variance of the Binomial distribution is given by *U*_*c*_*(t)* = *D(t)c*_*t*_*(1* − *c*_*t*_). For Raleigh, 23^2^ is added to the variance *U*_*c*_*(t)* to account for the (independent) uncertainty due to the aforementioned rounding of the case numbers, where 23 is the largest possible rounding error (*N/*20, 000).

The extended Kalman filter algorithm to estimate the state of the SEIR-WW system, based on different types of data, is presented in Algorithm 1. Our current implementation is done with custom MATLAB 2019b code; the process is however generally implementable in any programming language. For code references, see “Code availability” section. Given the update function *f(x)*, the observation matrices *C(t)*, the state noise *Q*, and the measurement error covariance *U(t)*, the method evaluates the state variables and their associated uncertainty matrix *P*.

The algorithm is used to calculate three different state estimates: 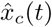 using only case number data (*C(t)*= *C*_*c*_*(t)*);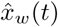 using only wastewater data *(C(t)* = *C*_*w*_ on days when wastewater sampling is done, otherwise Kalman update is skipped); 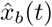 using both case and wastewater data *(C(t)* = *C*_*b*_*(t)* on days when wastewater sampling is done, *C(t)*= *C*_*c*_*(t)* otherwise). These were then used to estimate the data that were not employed for the state estimation, that is, we calculated 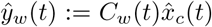 and 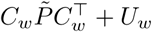. (*C*_*i*_ are the Kalman filter observation matrices, Eq. 6).

The Kalman filter is complemented with a simple outlier saturation for the wastewater data. The model-predicted value for a wastewater measurement is given by 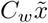, with prediction error variance 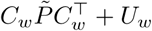. If the measurement differs from the model-prediction by more than four standard deviations, the measurement is replaced by the saturated value 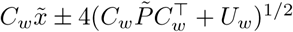.

#### Algorithm 1

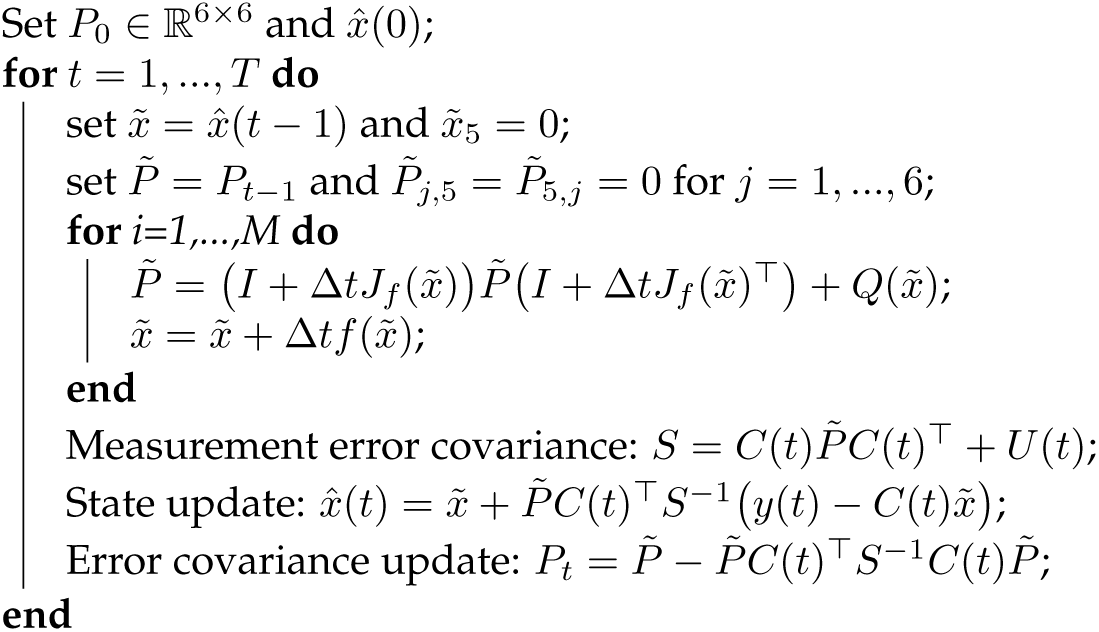

The Extended Kalman filter for the SEIR-WW model with time step Δ*t* = *1/M* (we use *M* = 10 d^*-*1^). *J*_*f*_ is the Jacobian of the function *f(x)*, obtained from the Jacobian of the reaction function by *J*_*f*_ = *BJ*_*r*_. The algorithm is standard, but the prediction step consists in solving a time-discretised ODE. The observation matrix *C(t)* is chosen from the three possibilities described in (6). Note the resetting of 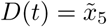 before the prediction loop.

### 1.4 Model parameters

As most time series data begin after the pandemic already diffused within a region, the initial sizes for the *E* and *I* compartments are directly automatically estimated from the data by

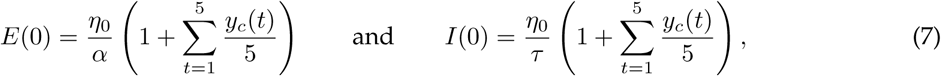

where *α* and *τ* are the transition rates *E* → *I* and *I* → *R*, respectively, whose inverses are the average duration an infected person remains in *E* and *I* compartments. *η*_*t*_ is the average ratio of total and detected cases at day *t*. Considering 5 data points is a trade-off between approximating values on the first day and sensitivity to noise. The model is little sensitive to this choice, *cf*. Supplementary Fig. 18. For Luxembourg, the data starts from the very beginning of the epidemic, when testing was not performed as actively as in the later stages. Therefore, we use *η*_*t*_ =3 for the first wave (until June 1, 2020), obtained from early prevalence studies [35]. Later, we use *η*_*t*_ = 1.8. This choice was cross-validated with an independent SEIR model fitted to Luxembourg data [25]. The reduction is partially due to the launch of a large scale testing campaign in Luxembourg [36], and partially to overall increased testing activity. For other regions, most available prevalence studies are only considering the early stages of the epidemic and are not usable for later stages. In the absence of additional reliable values, we maintain *η*_*t*_ = 1.8 for all other regions. It is possible to further calibrate such values with further tailored prevalence studies.

The daily ratios *c*_*t*_ of detected and total cases are obtained as follows. Initially, a weekly rhythm for case numbers is identified by averaging first over five weeks, and then by a moving average over three weeks:

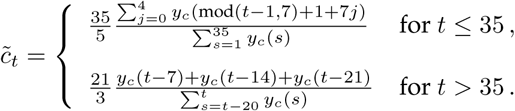

Then, these values are normalised by the weekly moving average:

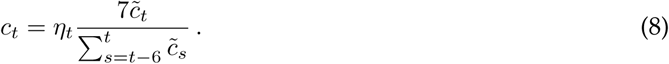

Note that the procedure for the first five weeks is not causal, but some data is anyway needed for model calibration. The later values *c*_*t*_ are causally determined from data. To obtain final values on public non-weekend holidays, *c*_*t*_ is reduced by a factor of 4 from the value given by Eq. 8 to account for reduced testing. In case the weekly rhythm is not regular, manual tuning could help improving performance (or estimating *c*_*t*_ based on number of performed tests, for example).

The variance of the wastewater measurements is estimated from data by

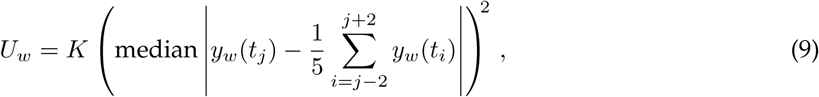

where each *t*_*i*_ is the time point when wastewater sampling is done. The scaling factor *K* is either 1*/*10 when wastewater data is used alone and *K* =1 when both case and wastewater data are used, as well as for the outlier detection. In the plots of wastewater data reconstruction, *K* = 1 is used for plotting the uncertainty envelope.

A final detail to consider when optimising the model to reproduce the observations: due to dilution, non-mixing environment and other factors, the dependency of the wastewater measurement on the number of detected cases is not perfectly linear [33] (see also Supplementary Fig. 1 and 2). Hence, we do a simple power transformation to the wastewater samples, for which the exponent *ε* is regarded as a tuning parameter of slight nonlinearity. *ε* and the other proportional parameters *γ* and *ν* are fitted by calculating the Kalman filter state estimate using the wastewater data, and then minimising the cost function

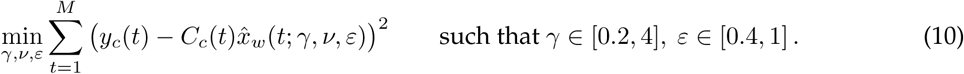

This way, we minimise the error in estimating the case numbers by the EKF state estimate using only wastewater data. Model parameters, either fixed by literature or fitted from Eq. 10, are reported in Supplementary Tab. 3.

The sensitivity of the model performance on assumed parameter values is assessed in Supplementary Fig. 18, which demonstrates the robustness of the model and justifies the current parameter choices. The sensitivity analysis was performed by varying the reference parameters up to *±* 50% of heir original value. The results are reported in Supplementary Fig. 18, using Luxembourg as a reference. For most parameters, the projections are consistent and slightly vary for values very far from the reference ones. The model is most sensitive to the parameter *c*_*t*_, which is usually estimated with independent methods. The minimal error corresponds to the reference value, while deviations induce larger errors. In our pipeline, changes in *c*_*t*_ are normally compensated by a change in *ν* by the same amount. This observation justifies the differing fitted values reported in Supplementary Tab. 4 for each region and recalls that, the more accurate seroprevalence studies are, the smaller the error associated with projections would be.

### 1.5 Analysis of model outputs

To obtain variables of epidemiological interest, we further analysed the state estimates outputted by the SEIR-EKF model. We recall that two estimates using only wastewater data are computed: one without interpolating data between sampling days (WW) and one with linear interpolation (ipWW).

The effective reproduction number *R*_eff_, the time-dependent average number of secondary infections from a single contagious case in a susceptible population [37], is directly extrapolated as [25]

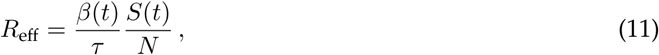

where *β* (*t*) and *S(t)* are state estimates. For *N* and *τ*, see Supplementary Tab. 3.

Short and mid-term projections are possible at any time *t*_*0*_ by stopping the Kalman filtering and simulating the model forward in time, starting from the latest state estimate *β(t*_*o*_) and keeping the infectivity parameter constant 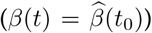. The effect of uncertainty in the parameter estimate *β*(*t*_*0*_) can be quantified by simulating envelopes using 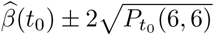 in the simulation (for every *t, P* (6, 6) represents the variance associated to *β* in the Kalman filter update, as discussed in Algorithm 1). Note that other uncertainties are omitted in these simulations; therefore, the short-term uncertainty in particular is under-estimated by the envelope. Projections based on case numbers alone, or on wastewater data, are consistent with each other within error bounds. Mid-term projection tests are reported in Supplementary Fig. 15 and discussed in Supplementary Note. As mid-term projections assume a constant *β* (*t*) over the time horizon, they mostly serve as a counterfactual analysis about the potential effects of social or pharmaceutical measures and/or changed viral infectivity. The large uncertainties reflect the set of potential changes of conditions.

Quantifying the quality of short-term projections using either case data only, wastewater data only, or both provides more reliable estimates of the epidemic unfolding over short time horizons. At each time step when wastewater data is available, the Kalman filter state estimation is stopped, and the SEIR-WW model is simulated *T* days forward without taking into account any new data. The total number of observed cases from the projection is calculated and compared with the actual number of observed cases during the same time horizon. Their absolute difference constitutes the prediction error. The prediction errors are standardised by the square root of the true number of cases, which represents the standard deviation estimate (assuming case numbers on a given time are binomially distributed). The standardised scores so obtained are then averaged over all time points on which the prediction is made, obtaining an overall average normalised error. To enable comparison between countries, the average standardised error is scaled per 100,000 equivalent inhabitants. Overall, the scaled average standardised prediction error *ξ* is:

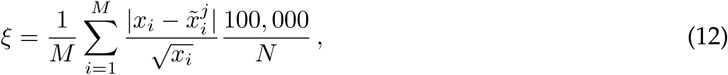

where *i* is the index of each point in any time horizon [*t*_0_,*T*] with *M* points in total; *j* is an index that considers the original type of data used for projections, i.e. *j* = {*c, w, b*} for case data only, wastewater data only, or both combined (note that, in the state estimate using combined data, the wastewater data are not interpolated); tilde-ed variables are the Kalman projections while non-tilde-ed variables correspond to measured data; *N* is the equivalent population of interest (*cf*. Supplementary Tab. 4).

## Notes

### Competing Interest Statement

The authors have declared no competing interest.

