## Supplementary material for "CoWWAn: Model-based assessment of COVID-19 epidemic dynamics by wastewater analysis"

### Supplementary Note: Mid-term projections

The extended Kalman Filter model (EKF) can be used to make short and mid-term projections, as described in the Main Text. These projections assume keeping the infectivity parameter constant ($\beta(t) = \beta(t_0)$) after any time $t_0$. Hence, as for all projections of this kind, their precision varies if they are conducted during a rapid increase of case numbers or during stable trends. These projections are usually employed to explore plausible scenarios, rather than for making forecasts. Hence, our interest lies in understanding their potential and benefits, and in proving that projections made with wastewater data are consistent with those made with case numbers. We list below several examples of mid- and long-term projections, starting from different time points. For illustrative purposes, we concentrate on Luxembourg, with combinations of wastewater and testing data.

Notably, making projections during stable or decreasing low-number periods might overlook unexpected outbreaks (Fig. 15a-b and g-h). Projections obtained when case numbers are high and increasing tend to overshoot on the long run (Fig. 15c-d) because of a constant $\beta$ corresponding to $R > 1$ (a reproductive number greater than 1 corresponds to diffusing epidemics). During stable trends, particularly if the forecasting horizon is not extremely long, the precision improves (Figs. 15e-f and i-j). These observations hold for both projections made with wastewater or testing cases: they are consistent with each other within error bounds, therefore making wastewater data a valuable resource for this kind of analysis as well. The large uncertainties reflect the set of potential changes of conditions: assumptions on the underlying SEIR model, large variability of social activities, non-pharmaceutical interventions being imposed, and so on. The EKF learns about epidemic changes only when new data are available, reflecting the effect of the various interventions. This calls for caution when interpreting mid-term estimates as plausible projections, rather than forecasts. On the other hand, mid- to long-term projections are valuable tools to analyse alternative possible scenarios that estimate the changes induced by different social measures or altered infectivity. This allows the comparison of different scenarios for epidemic management.

| Country | City | Population served | Units of measure | Source |
|---|---|---|---|---|
| Spain | Barcelona Prat de Llobregat | 2,000,000 | SARS-CoV-2 RNA copies/day | doi:10.5281/zenodo.4147073 https://sarsaigua.icra.cat/ |
| Canada | Kitchener | 242,000 | SARS-CoV-2 RNA copies /PMMoV copies | https://www.regionofwaterloo.ca/en/health-and-wellness/covid-19-wastewater-surveillance.aspx |
| Slovenia | Kranj | 40,000 | SARS-CoV-2 RNA copies/PMMoV copies | https://github.com/sledilnik/data |
| Switzerland | Lausanne | 240,000 | SARS-CoV-2 RNA copies/day/100,000 equivalent inhabitants | https://sensors-eawag.ch/sars/lausanne.html |
| Slovenia | Ljubljana | 280,000 | SARS-CoV-2 RNA copies/copies PMMoV | https://github.com/sledilnik/data |
| Luxembourg | Luxembourg | 610,000 | SARS-CoV-2 RNA copies/day/100,000 equivalent inhabitants | https://www.list.lu/en/covid-19/coronastep/ |
| USA | Milwaukee | 615,934 | MGC/person/day (gene copies per person per day) | https://www.dhs.wisconsin.gov/covid-19/wastewater.htm |
| Netherlands | Netherlands | 17,178,109 | SARS-CoV-2 RNA copies/day/100,000 equivalent inhabitants | https://data.rivm.nl/covid-19/COVID-19$_$rioolwaterdata.csv |
| USA | Oshkosh | 78,300 | MGC/person/day (gene copies per person per day) | https://www.dhs.wisconsin.gov/covid-19/wastewater.htm |
| USA | Raleigh | 460,000 | SARS-CoV-2 RNA copies/day/100,000 equivalent inhabitants | https://covid19.ncdhhs.gov/dashboard/wastewater-monitoring |
| Spain | Riera de la Bisbal | 100,000 | SARS-CoV-2 RNA copies/day | doi:10.5281/zenodo.4147073 https://sarsaigua.icra.cat/ |
| Switzerland | Zürich | 450,000 | SARS-CoV-2 RNA copies/day/100,000 equivalent inhabitants | https://sensors-eawag.ch/sars/zurich.html |

Supplementary table 1: **Considered regions, associated equivalent population served by sewage facilities (according to official sources), units of measure of wastewater data, and their sources.**

| Region | Method | 7-day window Error | SD | Difference | 14-day window Error | SD | Difference |
|---|---|---|---|---|---|---|---|
| Barcelona | Case data | 0.75 | 0.74 | | 1.62 | 2.05 | |
| | WW data | 1.17 | 0.99 | +55.7% | 1.97 | 1.90 | +21.9% |
| | WWip data | 1.10 | 0.84 | +46.6% | 1.71 | 1.67 | +5.4% |
| | All data | 0.69 | 0.64 | -7.6% | 1.52 | 1.77 | -6.2% |
| | Case data | 1.12 | 0.98 | | 2.30 | 2.27 | |
| Kitchener | WW data | 2.42 | 1.83 | +115.7% | 3.65 | 2.91 | +58.4% |
| | All data | 1.12 | 0.76 | +0.2% | 2.18 | 1.50 | -5.2% |
| (modified) | WW data | 1.51 | 0.96 | +34.2% | 2.68 | 1.61 | +16.3% |
| | All data | 1.04 | 0.81 | -7.2% | 2.12 | 1.66 | -7.8% |
| Kranj | Case data | 6.68 | 7.53 | | 13.2 | 24.0 | |
| | WW data | 11.8 | 7.02 | +76.0% | 17.0 | 10.6 | +28.6% |
| | WWip data | 11.2 | 7.58 | +67.6% | 16.3 | 11.2 | +23.2% |
| | All data | 6.43 | 6.09 | -3.8% | 12.2 | 18.6 | -7.3% |
| Lausanne | Case data | 1.59 | 1.57 | | 3.02 | 3.69 | |
| | WW data | 2.34 | 2.17 | +47.4% | 3.88 | 4.51 | +28.4% |
| | All data | 1.48 | 1.27 | -6.5% | 2.70 | 2.75 | -10.5% |
| Ljubljana | Case data | 1.66 | 1.99 | | 3.34 | 5.6 | |
| | WW data | 2.37 | 1.81 | +42.5% | 4.05 | 2.73 | +21.2% |
| | All data | 1.51 | 1.70 | -9.4% | 3.07 | 4.81 | -8.2% |
| Luxembourg | Case data | 1.26 | 1.38 | | 2.48 | 2.96 | |
| | WW data | 1.88 | 1.36 | +49.3% | 3.20 | 2.92 | +29.0% |
| | All data | 1.13 | 1.17 | -10.7% | 2.18 | 2.57 | -12.0% |
| Milwaukee | Case data | 0.84 | 0.74 | | 1.71 | 1.45 | |
| | WW data | 1.66 | 1.23 | +97.4% | 2.90 | 2.20 | +70.1% |
| | All data | 0.83 | 0.75 | -0.8% | 1.67 | 1.52 | -2.2% |
| Netherlands | Case data | 0.30 | 0.75 | | 0.92 | 3.40 | |
| | WW data | 0.44 | 0.29 | +46.3% | 0.74 | 0.60 | -19.3% |
| | All data | 0.29 | 0.73 | -4.1% | 0.87 | 3.21 | -5.0% |
| Oshkosh | Case data | 3.05 | 4.21 | | 5.19 | 7.57 | |
| | WW data | 10.4 | 7.01 | +241.5% | 14.2 | 10.1 | +174.2% |
| | All data | 3.06 | 3.54 | +0.6% | 4.63 | 5.75 | -10.8% |
| Raleigh | Case data | 1.66 | 1.77 | | 2.99 | 3.75 | |
| | WW data | 1.16 | 0.75 | -30.4% | 1.80 | 1.33 | -39.9% |
| | All data | 1.19 | 0.93 | -28.5% | 2.08 | 2.02 | -30.4% |
| Riera de la Bisbal | Case data | 3.11 | 2.60 | | 5.22 | 4.23 | |
| | WW data | 4.20 | 3.44 | +35.2% | 5.86 | 5.85 | +12.3% |
| | WWip data | 4.13 | 3.45 | +32.9% | 5.81 | 6.23 | +11.5% |
| | All data | 2.73 | 2.42 | -12.2% | 4.61 | 3.75 | -11.7% |
| Zurich | Case data | 1.02 | 0.86 | | 2.00 | 1.94 | |
| | WW data | 1.77 | 1.41 | +73.6% | 2.98 | 2.83 | +49.0% |
| | All data | 1.26 | 1.01 | +23.3% | 2.19 | 1.86 | +9.3% |

Supplementary table 2: **Summary results about short-term prediction performance, for different regions.** The table reports standardised and averaged prediction errors obtained from wastewater data alone or combined with case numbers, their standard deviations, and difference to benchmark (i.e.predictions based on case data). We consider either 7-day prediction window (left) or 14-day prediction window (right).
For Kitchener, the "modified" results are obtained by scaling the wastewater data after May 17, 2021 by a factor 0.4, as explained in Methods (Sec. Data). For three regions with low sampling frequency, results with interpolated wastewater data (WWip) are shown as well.
Projections made from wastewater data do not deviate much from those obtained from case numbers and are usually similar within error bounds. A notable exception is Oshkosh, for which the $c_t$ parameter during the beginning of 2021 should be updated to reflect undertesting.
Wastewater-based projections yield relatively smaller errors on longer time horizons, potentially due to their lower sensitivity to daily fluctuations (compare 7-days and 14-days window projections).
Combining wastewater and case numbers usually improves the prediction performance, suggesting the possibility to complement the testing routines with wastewater data, in order to obtain more reliable projections about future epidemic trends. This can be particularly useful on targeted areas with higher infectivity risk, to assess the efficacy of interventions.

| Symbol | Explanation | Value | Source |
|---|---|---|---|
| $\alpha$ | Rate $E \to I$ | 0.44 $d^{-1}$ | [1] |
| $\tau$ | Rate $I \to R$ | 0.32 $d^{-1}$ | [1] |
| $\beta(0)$ | Initial infectivity | 0.44 $d^{-1}$ | — |
| $\Delta t$ | Time step length | 0.1 $d$ | — |
| $q_{\beta,1}$ | Variance of $\beta(t+1) - \beta(t)$ when $t \leq 30$ | $0.05^2$ $d^{-2}$ | — |
| $q_{\beta,2}$ | Variance of $\beta(t+1) - \beta(t)$ when $t > 30$ | $0.005^2$ $d^{-2}$ | — |
| $\kappa$ | EKF sensitivity parameter | 4 | — |
| $N$ | Population size | regional | Sources in Tab. 1 |
| $\gamma$ | Rate $A \to \varnothing$ | regional | Fitted by Eq. (10) of Methods |
| $\nu$ | Ratio of $y_w$/A | regional | Fitted by Eq. (10) of Methods |
| $\varepsilon$ | Exponent in nonlinear mapping of WW data | regional | Fitted by Eq. (10) of Methods |
| $U_W$ | Measurement error variance of wastewater data | regional | Eq. (9) of Methods |
| $E(0)$ | Initial size of $E$-compartment | regional | Eq. (7) of Methods |
| $I(0)$ | Initial size of $I$-compartment | regional | Eq. (7) of Methods |
| var$(E(0))$ | Uncertainty of $E(0)$ | regional | $(E(0)/2)^2$ |
| var$(I(0))$ | Uncertainty of $I(0)$ | regional | $(I(0)/2)^2$ |

Supplementary table 3: **Model parameters: description** Parameter symbols, descriptions, values, and their sources. The parameter $q_\beta$, controlling the allowed change of $\beta(t)$ in one day, is changed after 30 days. This is done to allow rapid changes in the beginning of the pandemic, when a strict lockdown quickly suppressed its propagation and to account for errors in initial $\beta(0)$. $d$ stands for "days". When the source is not indicated, the parameter values is first initiated as an educated guess and then tested with sensitivity analysis (see Supplementary Fig. 18).
1 - Kemp, F. et al. Modelling COVID-19 dynamics and potential for herd immunity by vaccination in Austria, Luxembourg and Sweden. *J. Theo. Biol.* 110874 (2021).

| Parameter | Barcelona | Kitchener | Kranj | Lausanne | Ljubljana | Luxembourg |
|---|---|---|---|---|---|---|
| $N$ | 2,000,000 | 242,000 | 40,000 | 240,000 | 280,000 | 634,730 |
| $\gamma$ | 0.20 $d^{-1}$ | 4.00 $d^{-1}$ | 1.43 $d^{-1}$ | 3.05 $d^{-1}$ | 3.21 $d^{-1}$ | 1.62 $d^{-1}$ |
| $\nu$ | $4.86 \cdot 10^{-2}$ | 2.73 | 1.38 | $6.67 \cdot 10^{10}$ | 2.77 | $6.40 \cdot 10^{4}$ |
| $\varepsilon$ | 0.40 | 0.40 | 0.40 | 1.00 | 0.526 | 0.613 |
| $U_W$ | 656 | 2.68 | 58.5 | $3.43 \cdot 10^{23}$ | 511 | $1.75 \cdot 10^{12}$ |
| $E(0)$ | 1,824 | 379 | 17 | 156 | 76 | 8 |
| $I(0)$ | 2,527 | 525 | 24 | 203 | 105 | 11 |

| Parameter | Milwaukee | Netherlands | Oshkosh | Raleigh | Riera | Zurich |
|---|---|---|---|---|---|---|
| $N$ | 615,934 | 17,178,109 | 68,000 | 460,000 | 100,000 | 450,000 |
| $\gamma$ | 4.00 $d^{-1}$ | 0.368 $d^{-1}$ | 4.00 $d^{-1}$ | 4.00 $d^{-1}$ | 1.20 $d^{-1}$ | 0.547 $d^{-1}$ |
| $\nu$ | 0.134 | 185 | 3.73 | $2.58 \cdot 10^{3}$ | 12.7 | $1.87 \cdot 10^{7}$ |
| $\varepsilon$ | 1.00 | 0.500 | 0.866 | 0.434 | 0.400 | 0.789 |
| $U_W$ | 1.03 | $2.50 \cdot 10^{11}$ | 50.7 | $6.00 \cdot 10^{8}$ | $1.69 \cdot 10^{3}$ | $3.44 \cdot 10^{17}$ |
| $E(0)$ | 298 | 4,412 | 166 | 1,815 | 4 | 238 |
| $I(0)$ | 413 | 6,112 | 231 | 2,514 | 6 | 330 |

Supplementary table 4: **Model parameters: fitted values.** Region-dependent fitted parameter values. Initial values for SEIR compartments are in units of equivalent inhabitants.

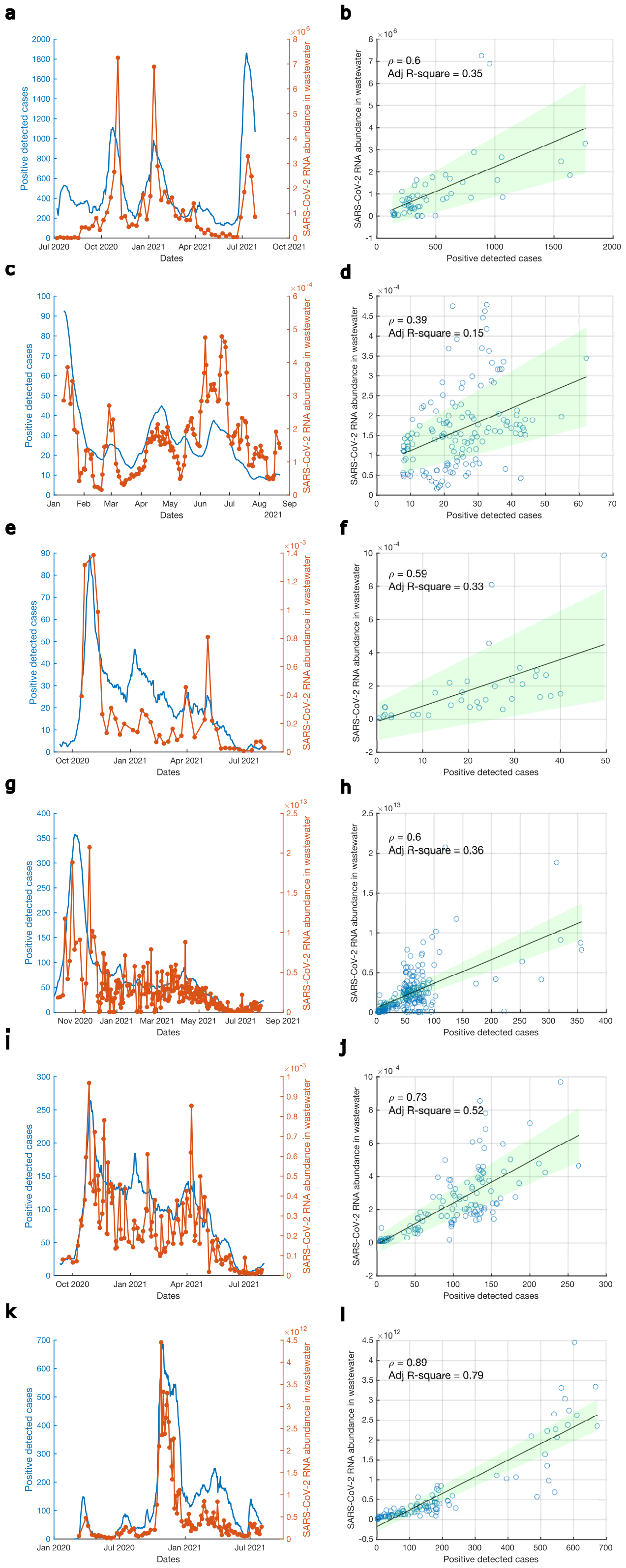


**Supplementary figure 1. Measured wastewater data and daily positive cases.** (a-b) Barcelona Prat de Llobregat, (c-d) Kitchener, (e-f) Kranj, (g-h) Lausanne, (i-j) Ljubljana, (k-l) Luxembourg. Left column: time series. Right column: wastewater data against daily positive cases: we observe a slight nonlinearity of cases vs wastewater samples, quantified by a Pearson's correlation $\rho < 1$ and by the adjusted R-square statistics for linear fit $< 1$ (both reported in the figure); the black line represents the linear fit, the green band is its $\pm 2\sigma$ error bound. Data sources and units of measures for wastewater data are reported in Supplementary Tab 1. Note that the reported correlation coefficients do not correspond to those reported in Fig. 2d of the main text: the latter represent the correlation of linear regression after data curation to reduce the noise, while the current ones are based on raw data.

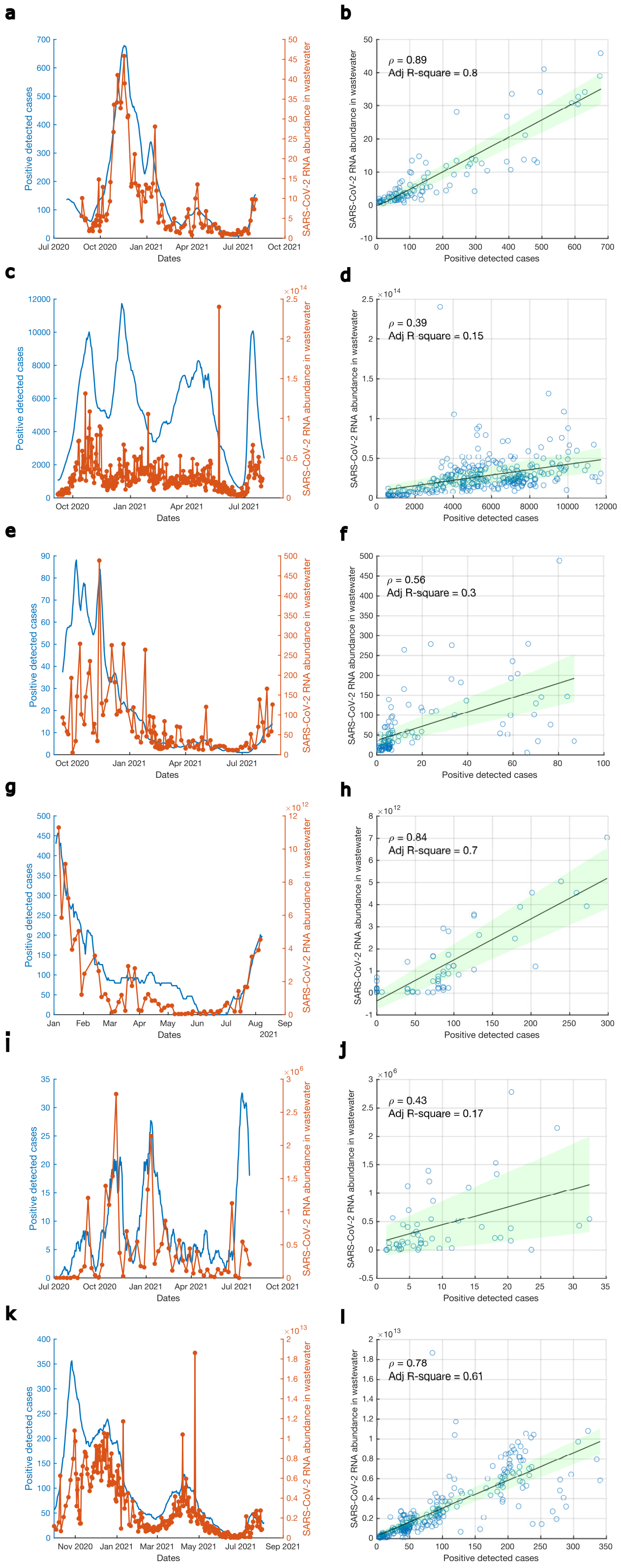


**Supplementary figure 2. Measured wastewater data and daily positive cases.** (a-b) Milwaukee, (c-d) the Netherlands, (e-f) Oshkosh, (g-h) Raleigh, (i-j) Riera de la Bisbal, (k-l) Zurich. Left column: time series. Right column: wastewater data against daily positive cases: we observe a slight nonlinearity of cases vs wastewater samples, quantified by a Pearson's correlation $\rho < 1$ and by the adjusted R-square statistics for linear fit $< 1$ (both reported in the figure); the black line represents the linear fit, the green band is its $\pm 2\sigma$ error bound. Data sources and units of measures for wastewater data are reported in Methods, Table 1. Note that the reported correlation coefficients do not correspond to those reported in Fig. 2d of main text: the latter represent the correlation of linear regression after data curation to reduce the noise, while the current ones are based on raw data.

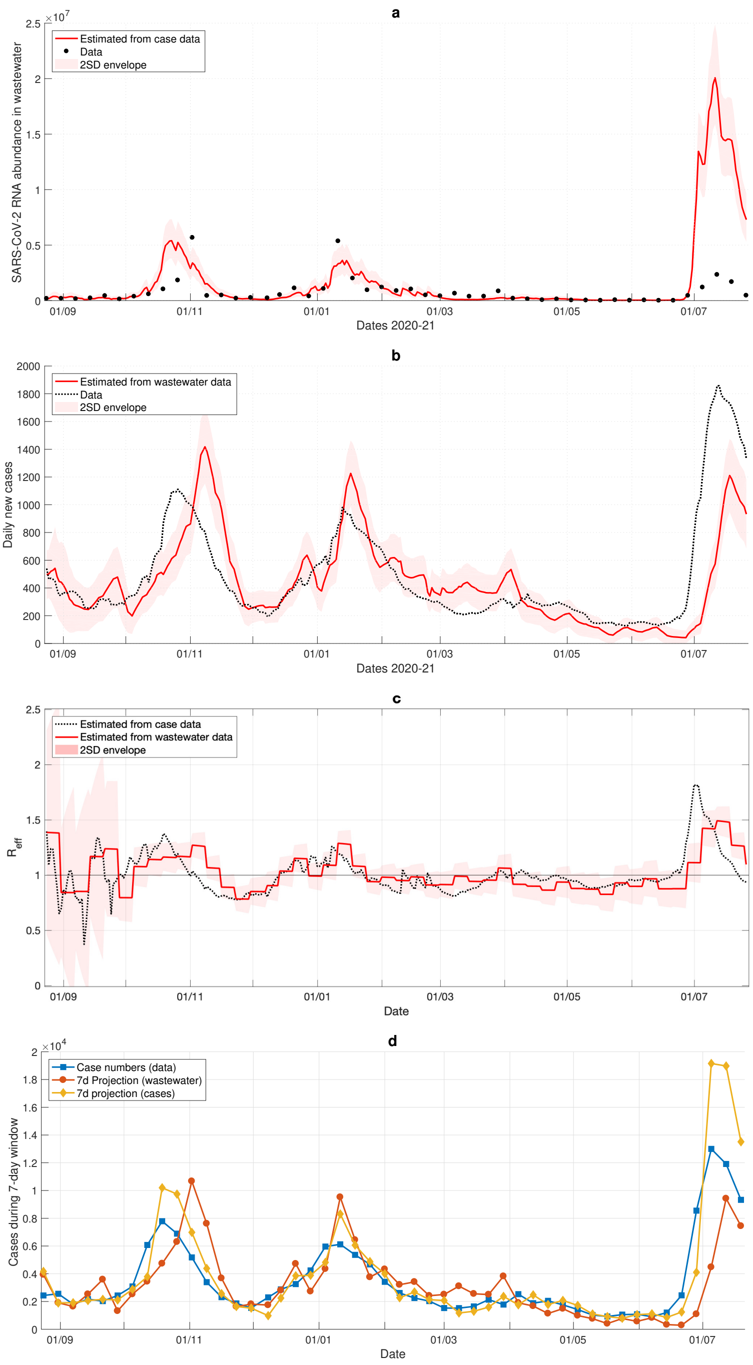


**Supplementary figure 3. Results for Barcelona Prat de Llobregat**. a: Reconstruction of wastewater data from case numbers. b: Reconstruction of case numbers from wastewater data. c: $R_{\text{eff}}$ estimated from case data and wastewater data. d: 7-day projections done at each day when wastewater sampling is done. The data shows number of cases in the 7-day time frame for which the projection is done.

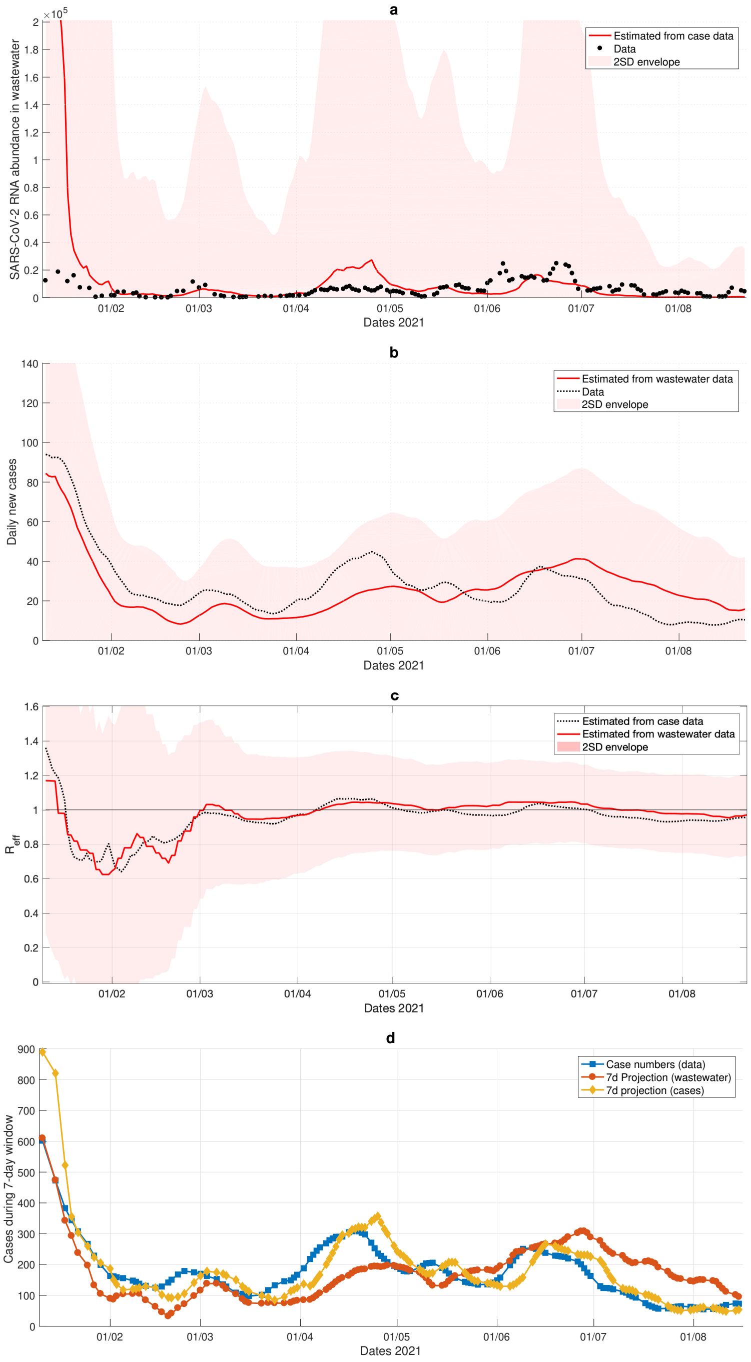


**Supplementary figure 4.** **Results for Kitchener**. a: Reconstruction of wastewater data from case numbers. b: Reconstruction of case numbers from wastewater data. c: $R_{\text{eff}}$ estimated from case data and wastewater data. d: 7-day projections done at each day when wastewater sampling is done. The data shows number of cases in the 7-day time frame for which the projection is done.

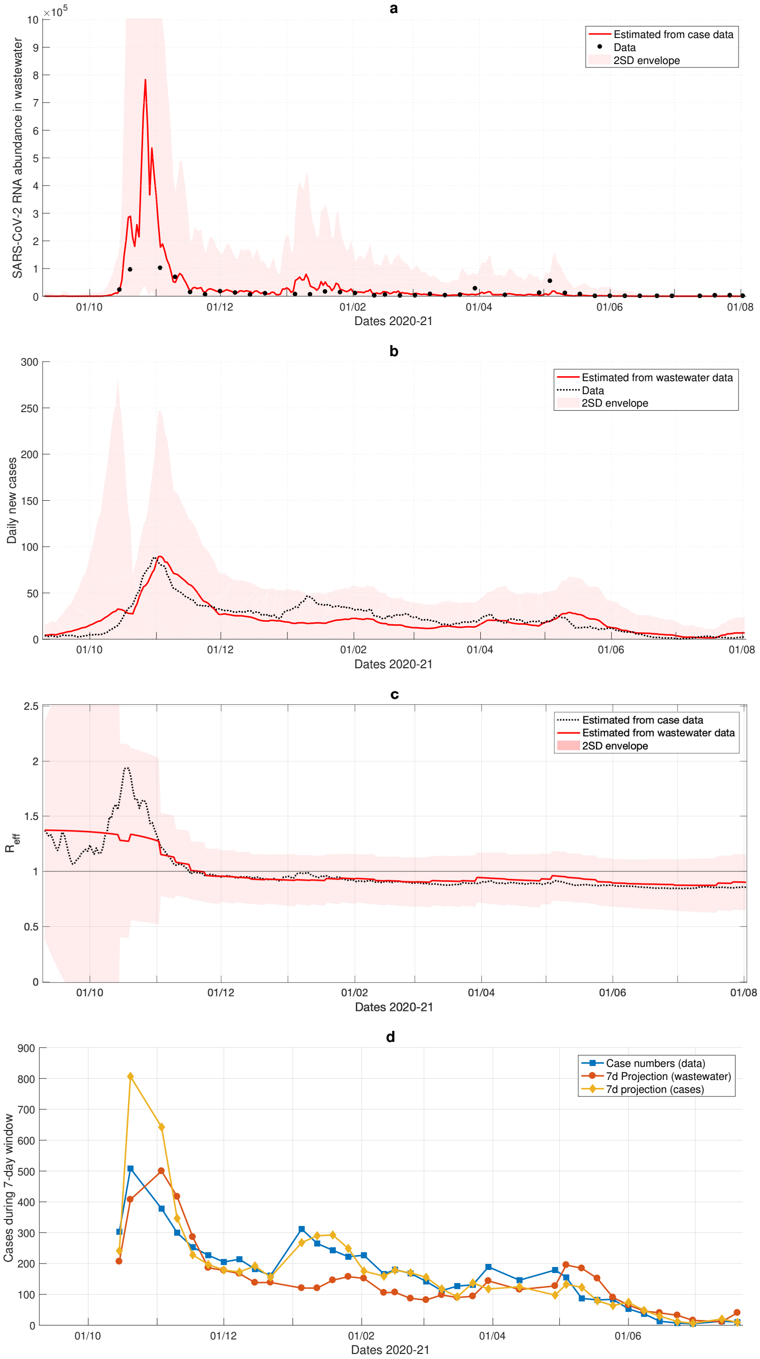


**Supplementary figure 5. Results for Kranj**. a: Reconstruction of wastewater data from case numbers. b: Reconstruction of case numbers from wastewater data. c: $R_{\text{eff}}$ estimated from case data and wastewater data. d: 7-day projections done at each day when wastewater sampling is done. The data shows number of cases in the 7-day time frame for which the projection is done.

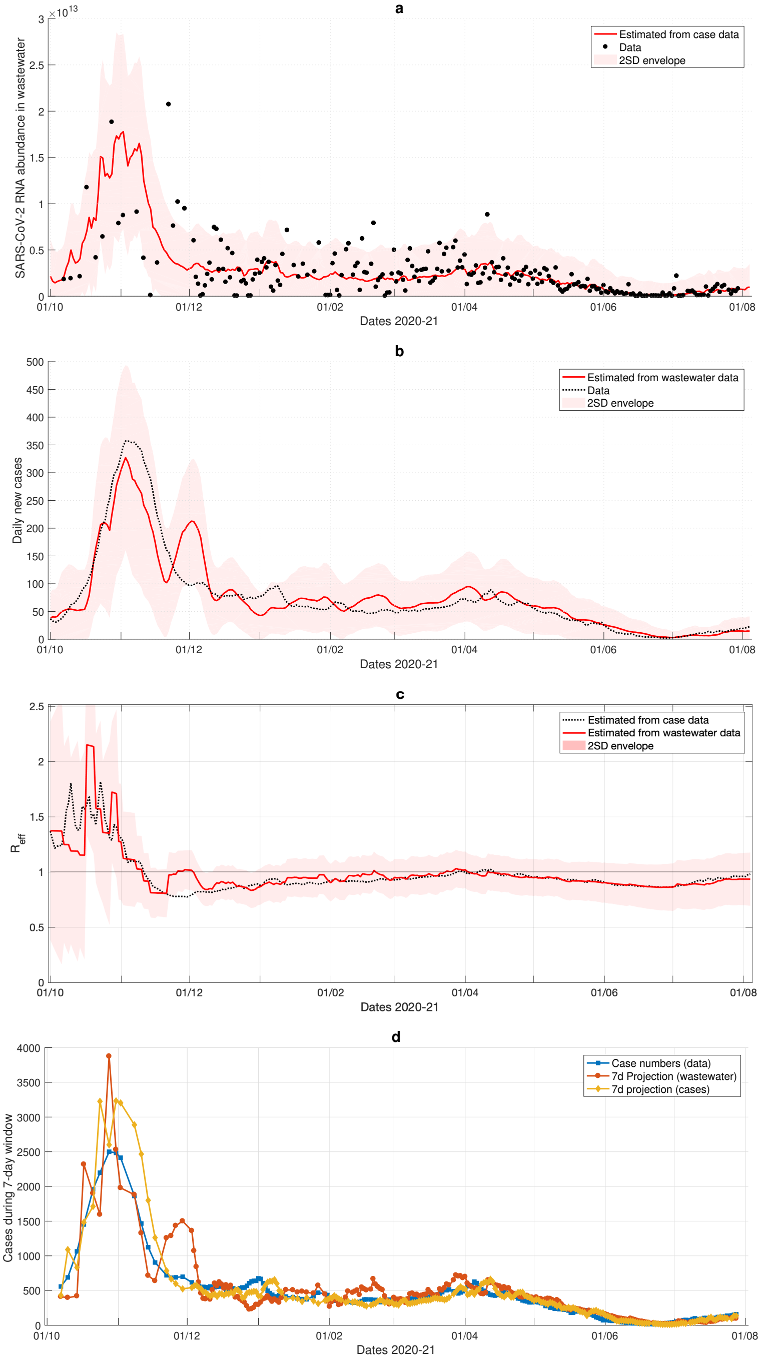


**Supplementary figure 6. Results for Lausanne**. a: Reconstruction of wastewater data from case numbers. b: Reconstruction of case numbers from wastewater data. c: $R_{\text{eff}}$ estimated from case data and wastewater data. d: 7-day projections done at each day when wastewater sampling is done. The data shows number of cases in the 7-day time frame for which the projection is done.

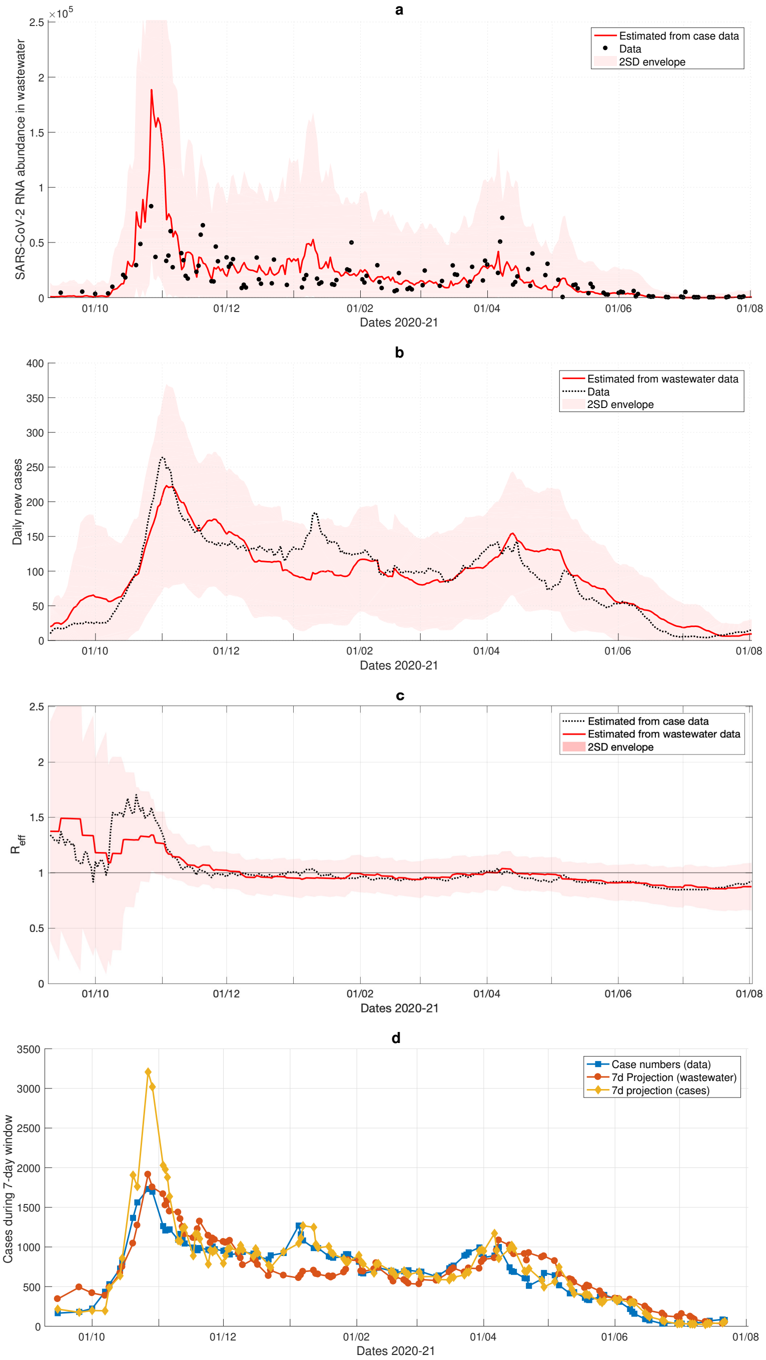


**Supplementary figure 7. Results for Ljubljana**. a: Reconstruction of wastewater data from case numbers. b: Reconstruction of case numbers from wastewater data. c: $R_{\text{eff}}$ estimated from case data and wastewater data. d: 7-day projections done at each day when wastewater sampling is done. The data shows number of cases in the 7-day time frame for which the projection is done.

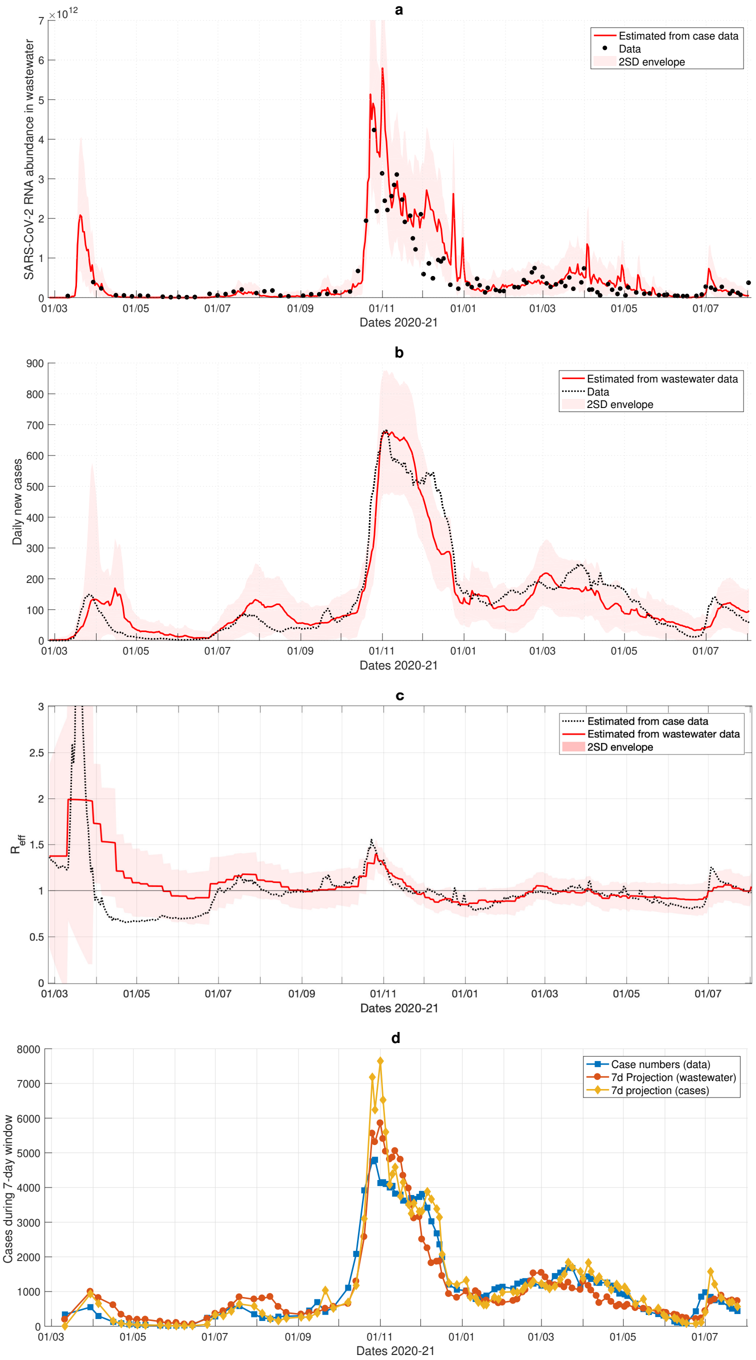


**Supplementary figure 8. Results for Luxembourg**. a: Reconstruction of wastewater data from case numbers. b: Reconstruction of case numbers from wastewater data. c: $R_{\text{eff}}$ estimated from case data and wastewater data. d: 7-day projections done at each day when wastewater sampling is done. The data shows number of cases in the 7-day time frame for which the projection is done.

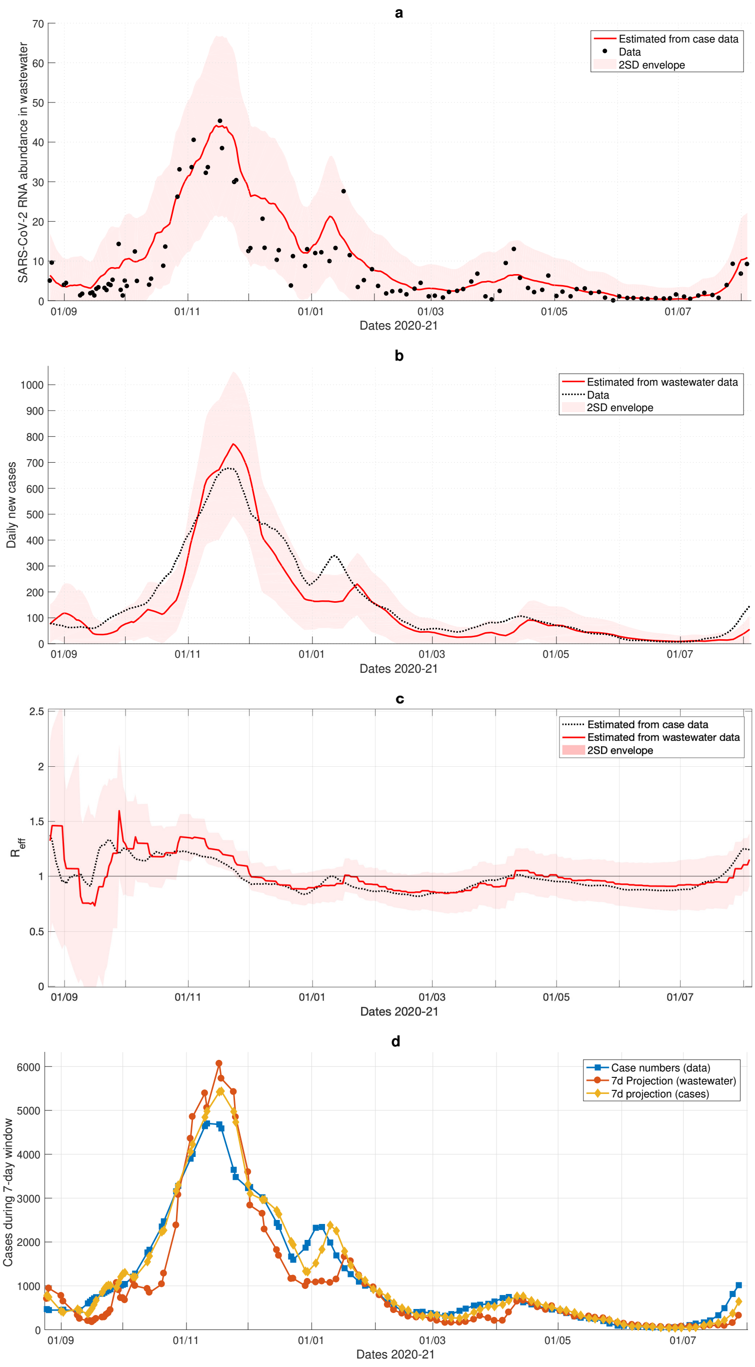


**Supplementary figure 9.** **Results for Milwaukee**. a: Reconstruction of wastewater data from case numbers. b: Reconstruction of case numbers from wastewater data. c: $R_{\text{eff}}$ estimated from case data and wastewater data. d: 7-day projections done at each day when wastewater sampling is done. The data shows number of cases in the 7-day time frame for which the projection is done.

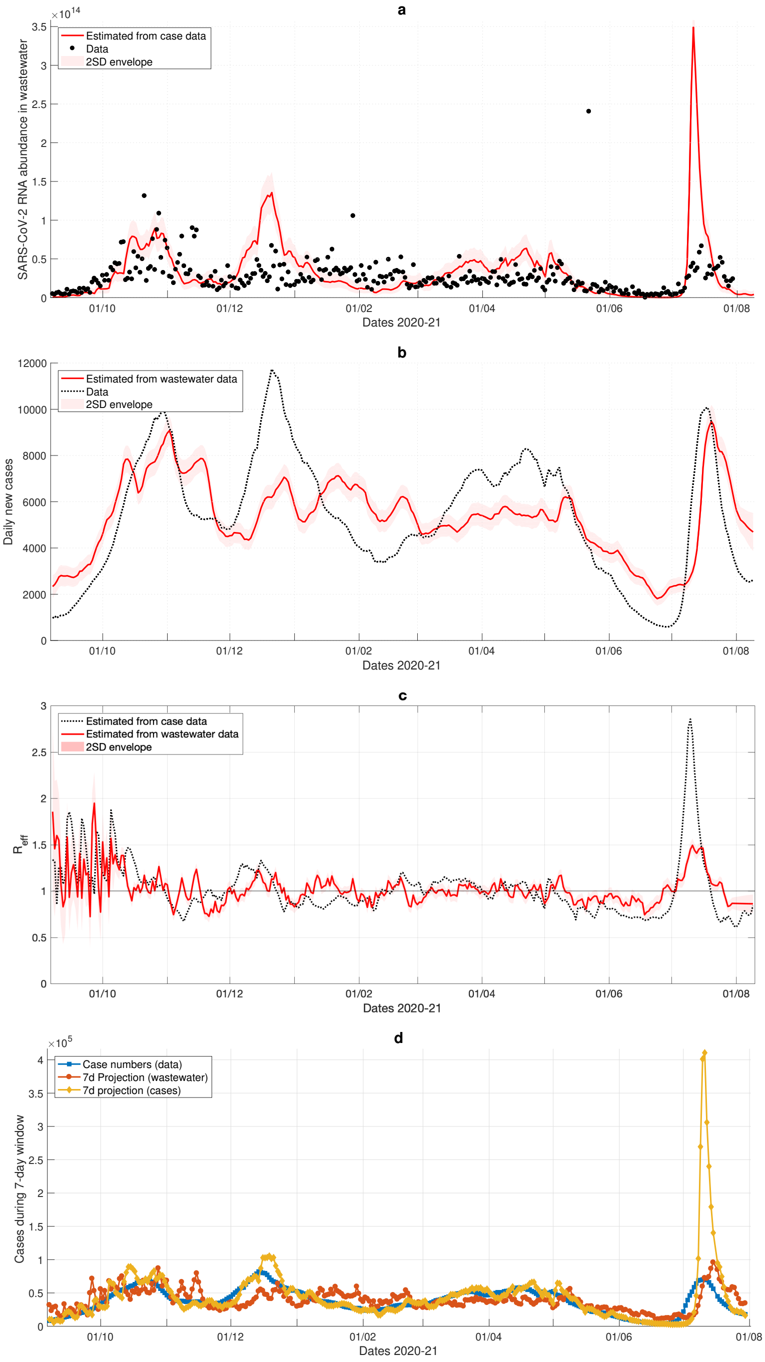


**Supplementary figure 10. Results for Netherlands**. a: Reconstruction of wastewater data from case numbers. b: Reconstruction of case numbers from wastewater data. c: $R_{\text{eff}}$ estimated from case data and wastewater data. d: 7-day projections done at each day when wastewater sampling is done. The data shows number of cases in the 7-day time frame for which the projection is done.

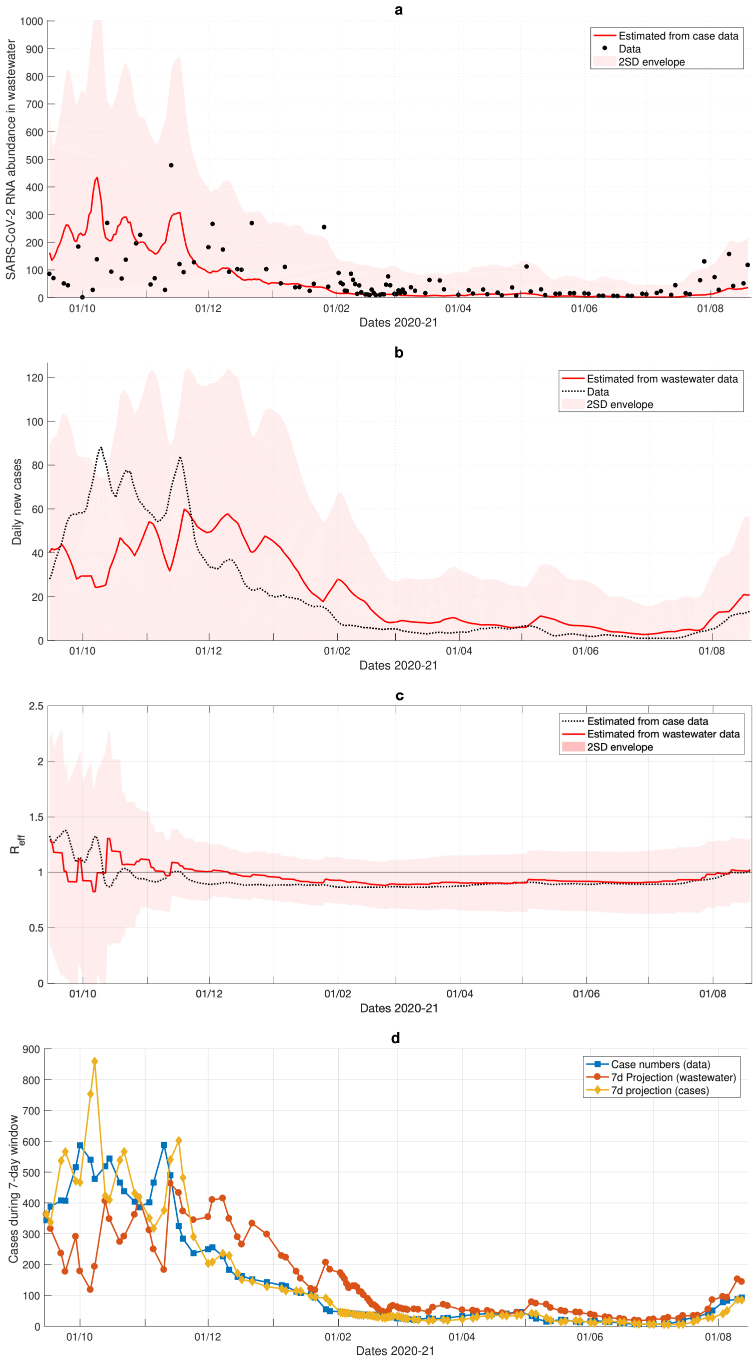


**Supplementary figure 11. Results for Oshkosh**. a: Reconstruction of wastewater data from case numbers. b: Reconstruction of case numbers from wastewater data. c: $R_{\text{eff}}$ estimated from case data and wastewater data. d: 7-day projections done at each day when wastewater sampling is done. The data shows number of cases in the 7-day time frame for which the projection is done.

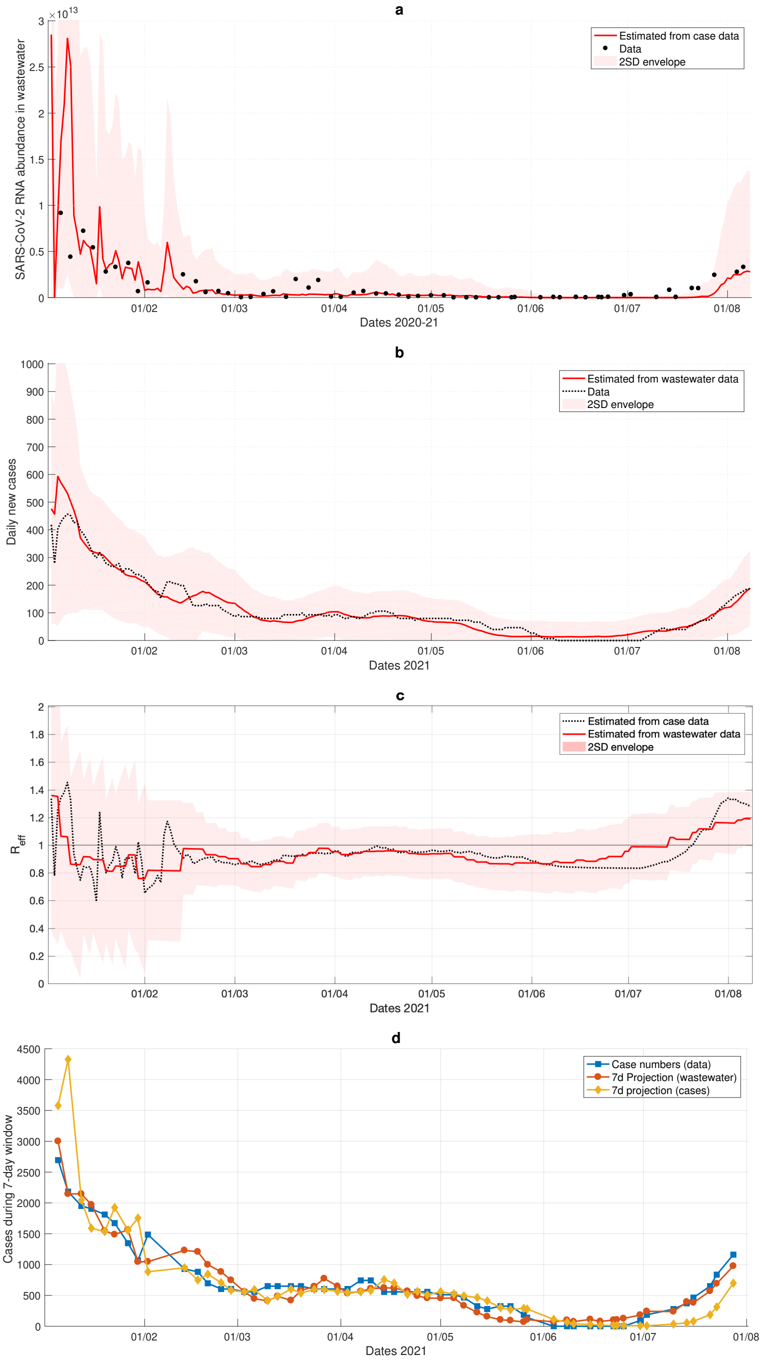


**Supplementary figure 12. Results for Raleigh**. a: Reconstruction of wastewater data from case numbers. b: Reconstruction of case numbers from wastewater data. c: $R_{\text{eff}}$ estimated from case data and wastewater data. d: 7-day projections done at each day when wastewater sampling is done. The data shows number of cases in the 7-day time frame for which the projection is done.

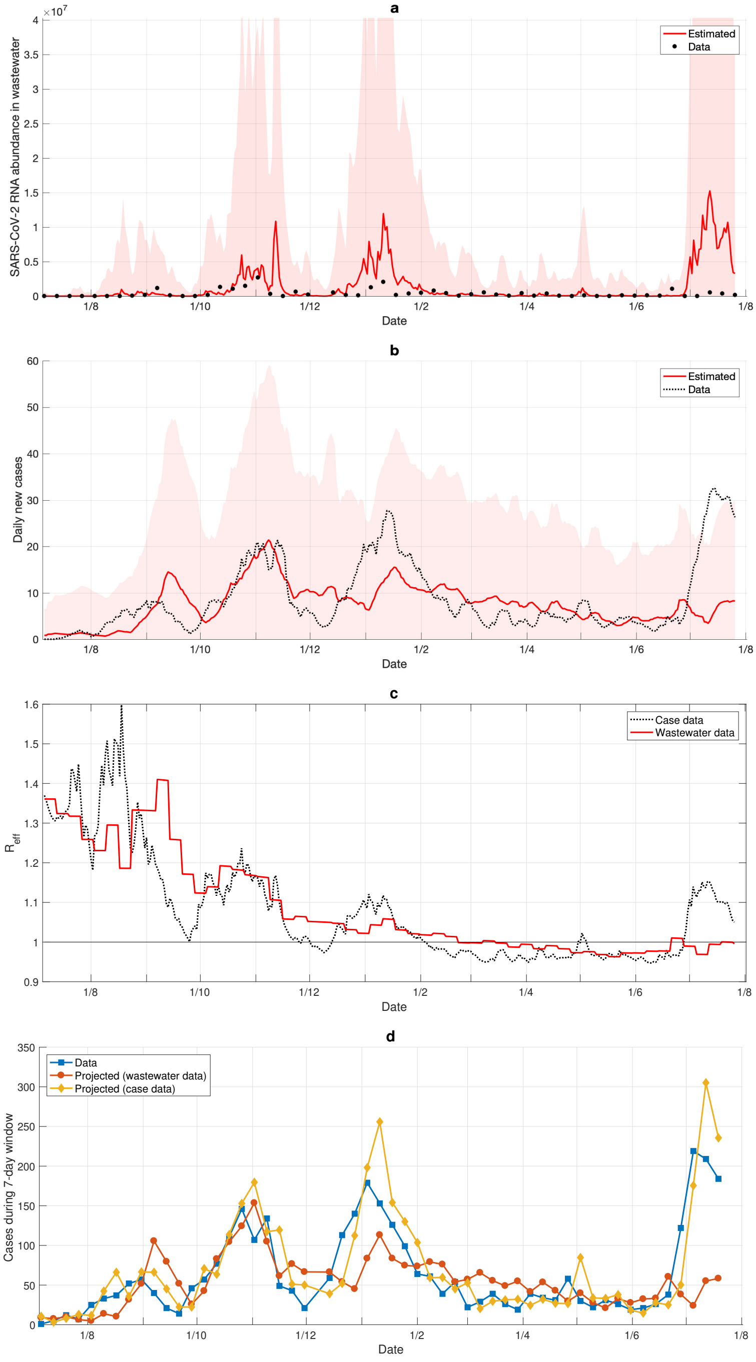


**Supplementary figure 13. Results for Riera de la Bisbal**. a: Reconstruction of wastewater data from case numbers. b: Reconstruction of case numbers from wastewater data. c: $R_{\text{eff}}$ estimated from case data and wastewater data. d: 7-day projections done at each day when wastewater sampling is done. The data shows number of cases in the 7-day time frame for which the projection is done.

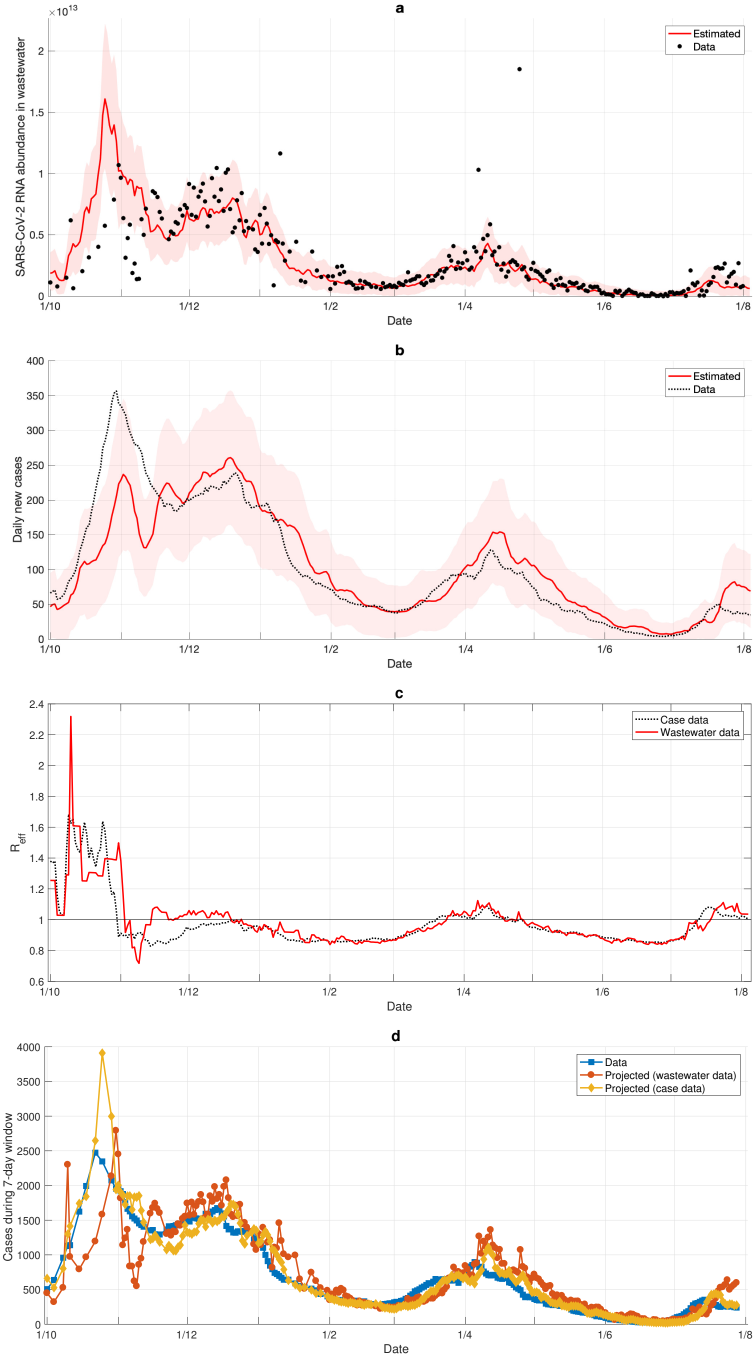


**Supplementary figure 14.** **Results for Zurich**. a: Reconstruction of wastewater data from case numbers. b: Reconstruction of case numbers from wastewater data. c: $R_{\text{eff}}$ estimated from case data and wastewater data. d: 7-day projections done at each day when wastewater sampling is done. The data shows number of cases in the 7-day time frame for which the projection is done.

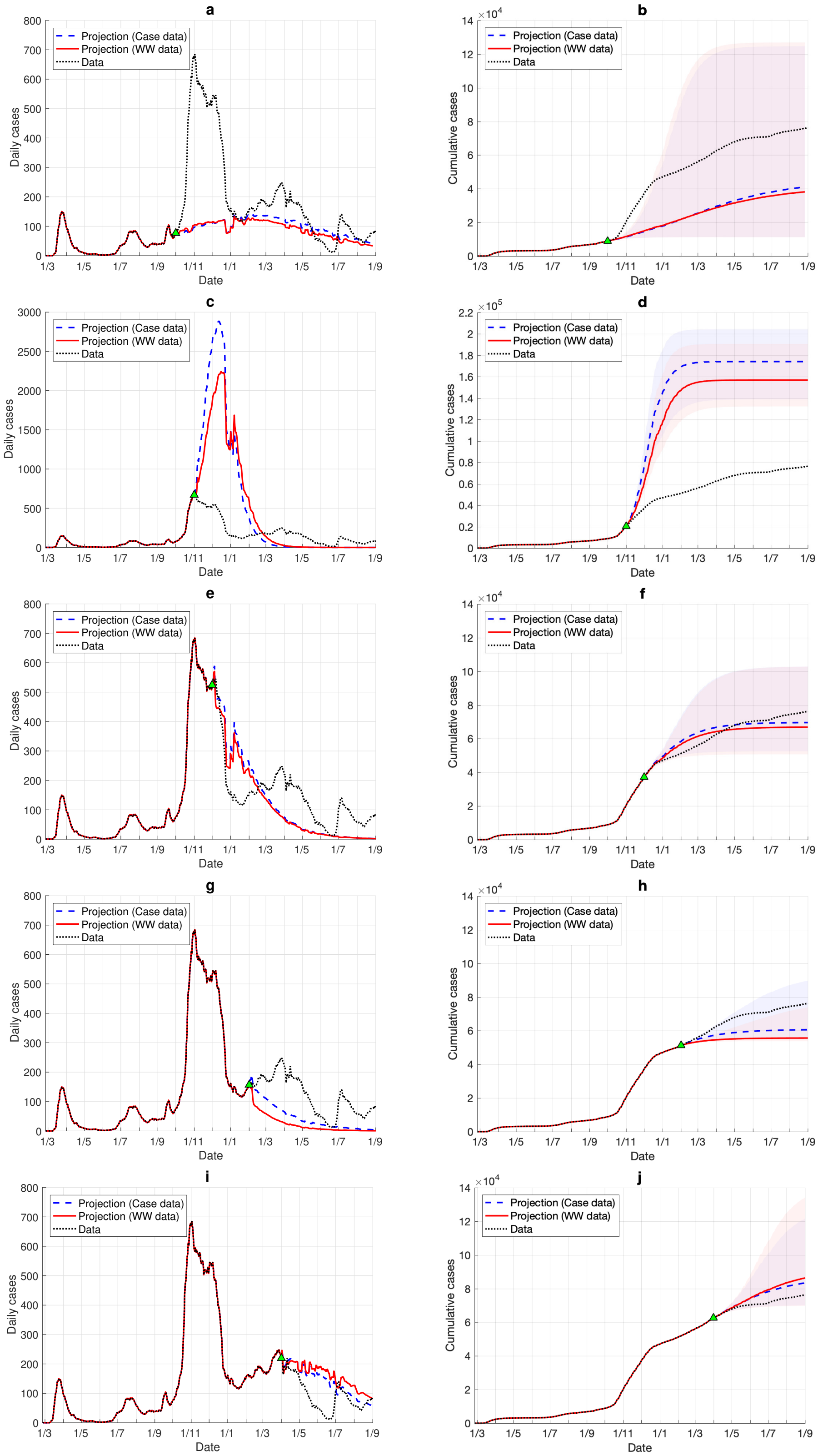


**Supplementary figure 15. Mid and long-term projections**. Start dates are: 01/10/2020 (a-b), 01/11/2020 (c-d), 01/12/2020 (e-f), 01/02/2021 (g-h), and 01/04/2021 (i-j). Start dates are indicated by a green triangle. The left column shows infection curves of daily cases, the right column of cumulative cases. Blue and red ribbons represent $\pm 2\sigma$ error bounds ($\sigma$ corresponds to a standard deviation). Note that the ribbons might overlap. All examples refer to Luxembourg data.

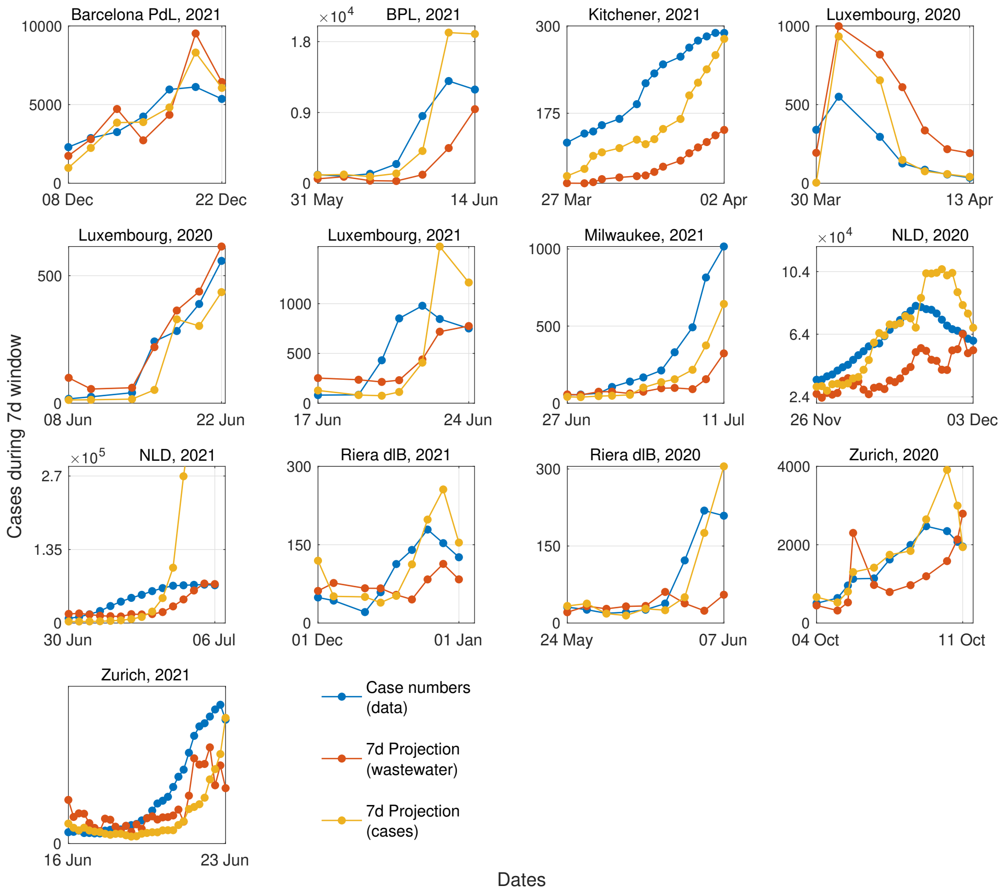


**Supplementary figure 16. Zoom into the epidemic resurgences visually recognised in the considered regions**. The figure displays short-term projections, made from wastewater data or case numbers, as well as true data for the 7-day time frame for which the projection is done. This figure completes Fig. 2d of main text with other epidemic resurgences observed in the considered regional areas. In general, consistent and noise-aware increasing trends need to be observed for some days before an online detection system can trigger a reliable alert. In addition, the wastewater sampling frequency plays a role in tuning the lead time of the warning. Hence, we do not often observe very advanced early warnings (as one might infer from retrospective observations like in Supplementary Fig 1 and 2), but we quantitatively verify the need for cautious interpretation.

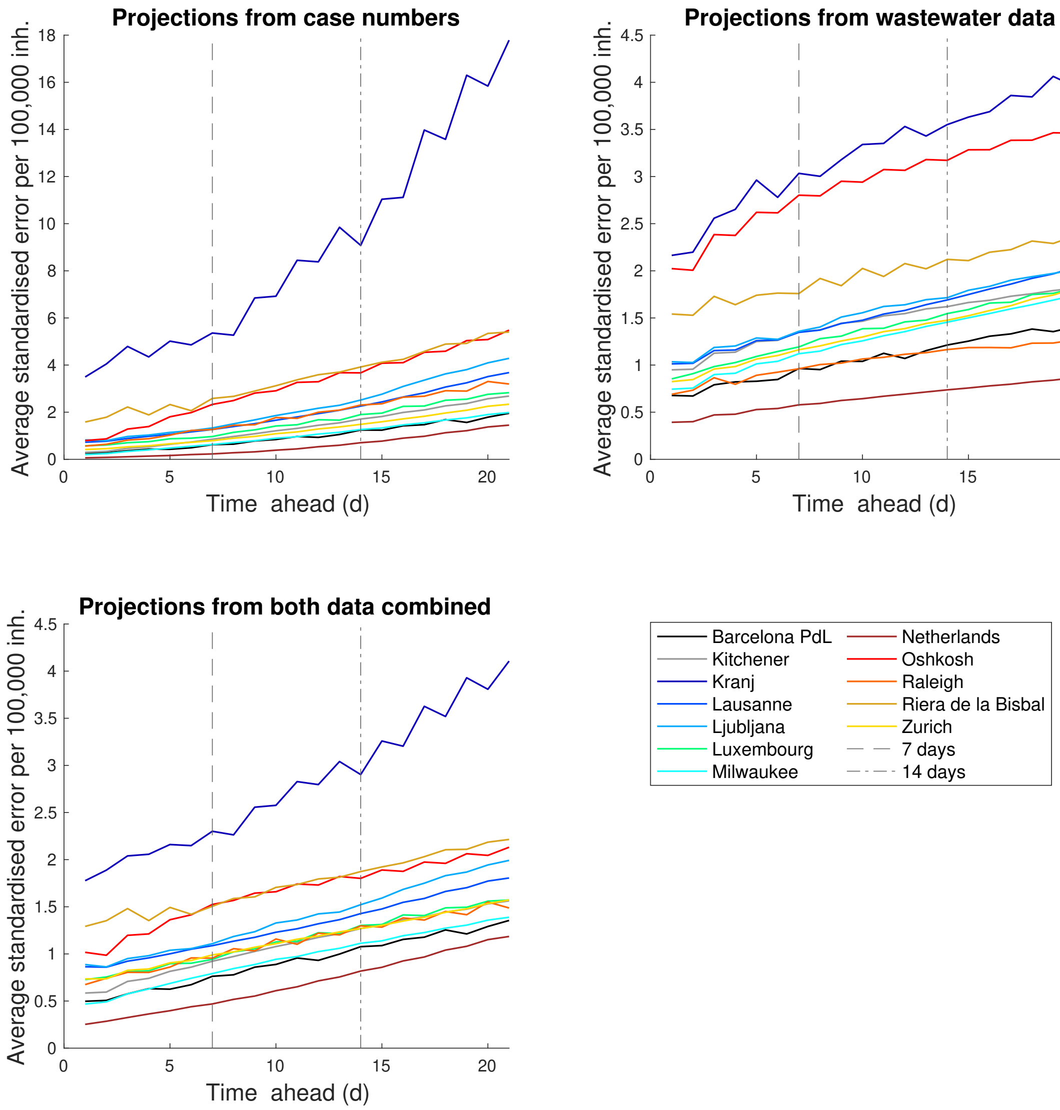


**Supplementary figure 17. Average standardised error vs projection time frame, for all countries**. The three panels refer to projections made from case numbers, from wastewater data, or from both data combined. Their mean values and 80% confidence intervals are reported in Fig. 2 of main text. Values at 7 and 14 days correspond to the ones reported in Supplementary Tab. 2.

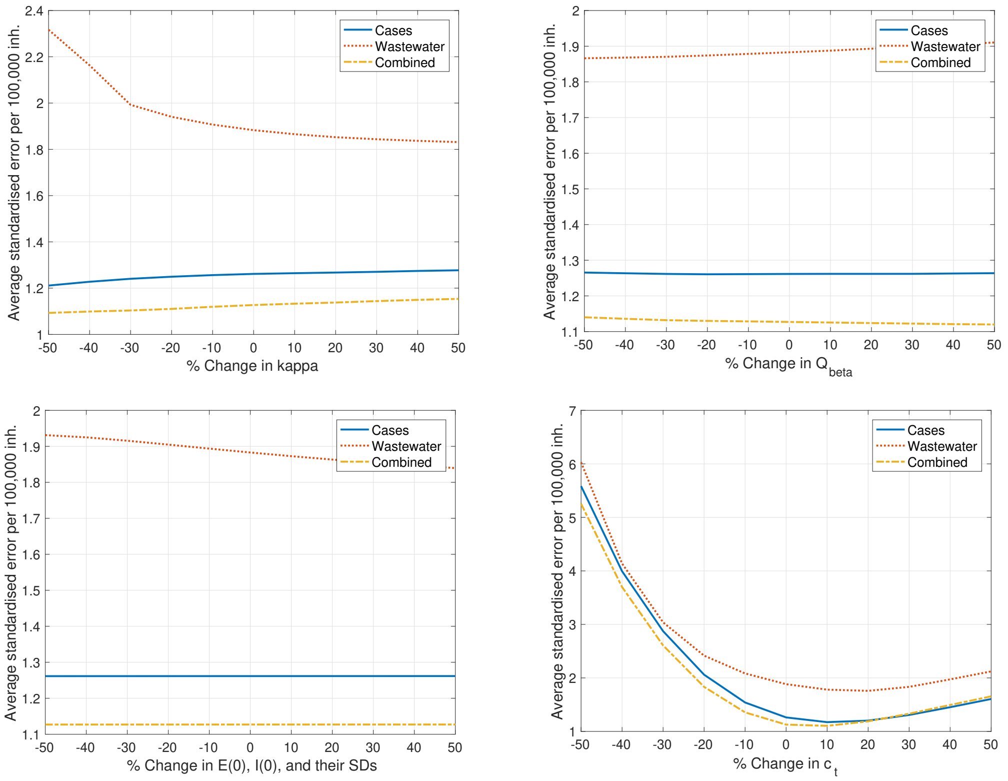


**Supplementary figure 18. Sensitivity analysis against changes in hand-picked parameters.** The plots show the changes in 7-day window prediction performance for Luxembourg when a parameter is changed. The no-change numbers (0%) are those shown for Luxembourg in Supplementary Tab. 2. The "normalised error" corresponds to the measure employed throughout the main text and defined in Eq. 10 of Methods. Change in $c_t$ (the share of detected cases) is compensated by a change in $\nu$ by the same amount. Note that other parameters are kept fixed. Some of the change could be compensated by re-running the parameter tuning pipeline.
